# Genome-wide association study reveals two novel genetic loci associated with chronic lung allograft dysfunction

**DOI:** 10.1101/2025.11.05.25339596

**Authors:** Simon Brocard, Vincent Mauduit, Martin Morin, Léo Boussamet, Nayane dos Santos Brito Silva, Axelle Durand, Pierre Halitim, Benjamin Renaud-Picard, Benjamin Coiffard, Xavier Demant, Loïc Falque, Jérome Le Pavec, Antoine Roux, Thomas Villeneuve, Christiane Knoop, Claire Merveilleux, Mathilde Salpin, Nicolas Carlier, Pierre Antoine Gourraud, Antoine Magnan, David Lair, Laureline Berthelot, Mario Südholt, Nicolas Vince, Adrien Tissot, Sophie Limou, COLT consortium

## Abstract

**Background:** Chronic lung allograft dysfunction (CLAD) leads to declining respiratory function and high mortality, representing the main barrier to long-term survival in lung transplantation (LT). We performed the first genome-wide association study (GWAS) investigating donor’s and recipient’s genetic factors associated with CLAD.

**Method:** We genotyped 392 donor-recipient pairs from the multicentric Cohort in Lung Transplantation. We tested 4.5 million SNPs for association with CLAD using multivariable logistic regression models corrected for age, sex, initial disease and genetic ancestry. Three levels of explanatory variables were separately considered to conduct GWAS: donors-only, recipients-only, and donor-recipient mismatches. We also ran HLA-centric analyses using the same models.

**Results:** Our analysis confirmed the deleterious impact of *HLA* allelic and epitopic mismatches on CLAD risk, mostly driven by class I HLA (p=0.004). No significant associations with CLAD were found for donors’ genotypes or donor-recipient non-HLA mismatches. We highlighted two independent recipient’s loci associated with CLAD, including one protective signal (0.39 in CLAD vs 0.66 in non-CLAD recipients, p-value=5.05×10^-7^, q-value=0.017, OR=0.35) encompassing the *PLXDC2* gene, and one risk signal (0.66 in CLAD vs 0.38 in non-CLAD recipients, p-value=9.86×10^-7^, q-value=0.017, OR=2.83) encompassing the *ZNF518A/BLNK* genes. These non-coding SNPs are putative regulatory variants of gene expression. Importantly, our single-cell RNA-sequencing showed a down-regulation of *PLXDC2* in monocytes and lung epithelium in CLAD vs healthy controls (p≤2.0×10^-16^).

**Conclusion:** This first LT GWAS revealed two candidate loci from the recipient’s genome, both biologically relevant for CLAD pathogenesis. Our study calls for larger LT genomic initiatives to increase power for signal discovery.

## Introduction

The number of lung transplants has risen steadily over the years. In 1990, around 250 lung transplants were performed worldwide compared to over 4,000 per year nowadays(1). The long-term survival rate remains around 63% in Europe after 5 years post-transplantation(2). Chronic lung allograft dysfunction (CLAD), defined by a persistent decline of at least 20% in FEV1 (forced expiratory volume in 1 second) from baseline, is the primary factor limiting long-term survival of lung transplants(3). Numerous risk factors for CLAD have been identified, including alloimmune (acute cellular or antibody-mediated rejection) and non-alloimmune factors (primary graft dysfunction, pulmonary infections, air pollution or gastro-esophageal reflux disease)(3,4). Several studies have explored the relationship between cytokine or other inflammation-related candidate genes’ polymorphisms and CLAD following lung transplantation (LT). First, *IFNG* and *TGFB1* polymorphisms were associated with graft fibrosis, a hallmark of chronic rejection(5). Later, *IFNG* and *IL6* genetic variants were associated with earlier CLAD onset, while no link was established with *TNF, TGFB1*, or *IL10* polymorphisms(6). However, these different findings were not replicated in subsequent studies and had a limited statistical power due to limited sample sizes (<100 patients)(7,8). Unlike renal transplantation, *HLA* matching between donors and recipients is usually not performed in LT(9). Large registry studies have repeatedly demonstrated an association of HLA mismatches (mostly defined as A-B-DRB1 mismatches) with patient and graft survival(10–12). Several reports also highlighted an association between *HLA* matching and CLAD subtypes, but it seems less clear whether the associations are driven by *HLA* class I-only, *HLA* class II-only or total HLA mismatches(11,13). In a recent single-center study, *HLA-DQA1/DQB1* epitopic mismatches were associated with an increased risk of CLAD(14). These findings underscore the complex genetic underpinnings of CLAD and the need for deeper investigations(13). Here, we performed an HLA-centric association analysis and the first non-*HLA* genome-wide association study (GWAS) investigating donor’s genetic factors, recipient’s genetic factors and donor-recipient mismatches associated with CLAD.

## Materials and Methods

### GenCOLT biobank

The GenCOLT cohort(15) is an ancillary study of the COLT (Cohort in Lung Transplantation) project(16). To date, GenCOLT has collected DNA samples from 392 pairs of lung transplant donors and recipients (total of 784 individuals) across 12 centers, as described in the Supplementary methods and in our previous publication(15). We selected case participants without distinction between the different CLAD subtypes, and controls were defined as patients without CLAD and with stable lung function at 5 years post-LT.

### SNP genotyping quality control, imputation and mismatch calculation

Genotyping quality control and SNP-to-SNP imputation were performed using gold standard recommendations and are further described in the Supplementary methods and in our previous report(15). We defined genetic ancestry percentages per individual with a detailed breakdown within five major ancestral groups (African, American, East Asian, European and South Asian) using <u>Admixture</u> (v.1.3.0)(15). Donors and recipients with more than 80% estimated European ancestry were classified as European(15) and kept for subsequent statistical analyses. We investigated both coding and non-coding mismatches in order to capture the possible immunogenic antigens, as well as the possible regulatory variants impacting the expression level of proteins important for alloimmune responses or graft injury. We defined SNP mismatches between the donor and the recipient from each pair for every high-quality SNP by considering whether the donor was bringing new allele(s) to the recipient and encoded the mismatches under an additive model (0, 1 or 2 according to the number of differences/mismatched nucleotides).

After these steps, a total of 7.3 million imputed SNPs and 67 European CLAD cases versus 124 European controls were available for the recipients’ analysis, 67 cases versus 123 controls for the donors’ analysis, and 64 cases versus 117 controls for the donor-recipient mismatches’ analysis (Fig.1).

**Figure 1:**
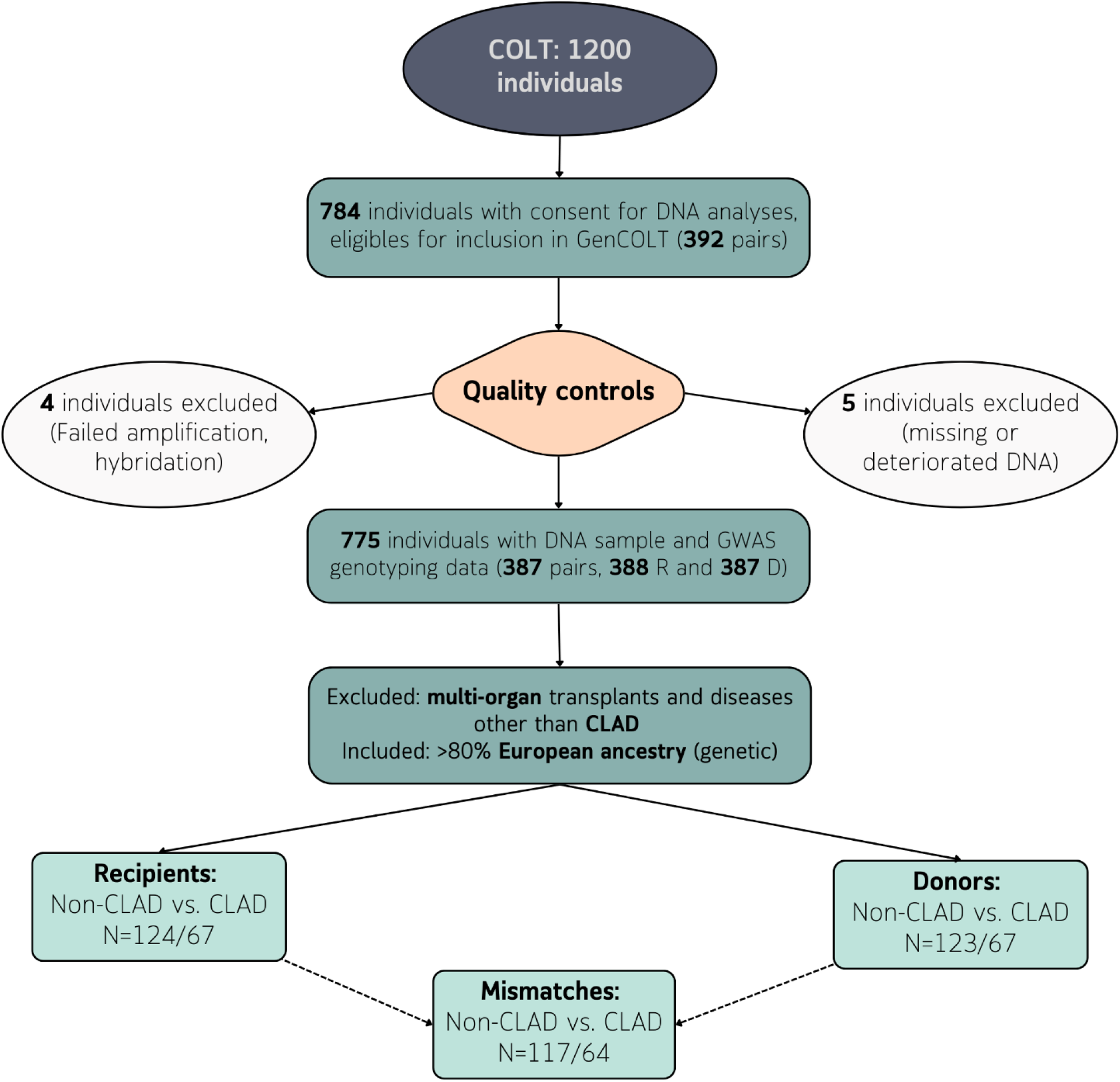
Flowchart of data processing upstream the genomic analyses. After quality controls for DNA, genotyping quality, 64 CLAD and 117 Non-CLAD patients were selected for the GWAS analysis in the mismatch setting, 67 CLAD vs 123 non-CLAD in the donor only setting and 67 CLAD vs 124 non-CLAD in the recipient only setting. CLAD: chronic allograft dysfunction, GWAS: genome-wide association study

### Imputation of *HLA* genotypes, allelic and epitopic mismatch calculation

To specifically assess the classical *HLA* polymorphic genetic region, SNP data were extracted from the major histocompatibility complex locus (chromosome 6 from 29 to 34 Mb). Following the SNP-HLA Reference Consortium (SHLARC) guidelines(17), two-field *HLA* genotype imputation was performed from SNP data for a total of 5 class I and II HLA genes (*HLA-A, HLA-B, HLA-C, HLA-DRB1* and *HLA-DQB1*) with HIBAG(18) and the reference panel consisting of multiethnic samples from the 1000 Genomes Project(19). In addition, we computed the HLA allelic mismatch scores capturing the donor’s and recipient’s allele differences at each *HLA* genetic locus (0, 1 or 2), within class-I *HLA* genes (HLA-A, HLA-B and *HLA-C*; score 0-6), class-II *HLA* genes (*HLA-DRB1* and *HLA-DQB1*; score 0-4), and both class I and II *HLA* genes (total *HLA* allelic mismatches; score 0-10). Finally, *HLA* epitopic mismatches were computed using the HLA-Epi software(20) available from the <u>EasyHLA</u> web platform. They were defined as the sum of eplet mismatches across *HLA* class I-only genes, *HLA* class II-only genes, and both *HLA* class I and II genes (total epitopic mismatches).

### Statistical analysis

#### Demographic and phenotype data description

Classical descriptive statistics such as mean, standard deviation and percentage were used to summarize patient characteristics using R (v.4.4.2) and the *gtsummary* (v.2.0.4) and *flextable* (v.0.9.7) libraries. Differences between patients with and without CLAD were assessed using the Wilcoxon rank-sum test for continuous variables, and either Fisher’s exact test or Pearson’s chi-square test for categorical variables, depending on expected counts.

#### *HLA* and SNP *association testing*

The HLA-centric analysis and GWAS were performed under an additive model of inheritance to capture all common (frequency >2% for HLA and >10% for GWAS) genetic determinants of CLAD, at three levels: (1) donors-only, (2) recipients-only, and (3) donor-recipient mismatches (Fig.S1). Further details on model construction and multiple testing corrections are provided in the Supplementary methods.

#### Top signals genetic and in-silico analyses

To further analyze the top signals, we used Cox proportional hazard models fitted with time-to-CLAD adjusted for the same covariates as the logistic regression models. The proportional hazards assumption was checked using the R Survival package (v.3.7.0). Survival curves were computed through the Kaplan-Meier method, with right-censoring applied for individuals who were event-free or lost to follow-up at the latest available timepoint. Differences in survival rate between groups were compared using a log-rank test.

We investigated the potential selective pressure signatures by retrieving the fixation index statistics (F_ST_) calculated two-by-two between each continental population from the 3DSNP data(21), and computing the integrated haplotype homozygosity score (iHS) with rehh (v.3.2.3).

Finally, to support the genetic and immunogenetic analyses, several publicly available databases and resources were explored. The GTEx (Genotype-Tissue Expression) project was used to assess tissue-specific gene expression and expression quantitative trait loci (eQTLs)(22). PROMAD, a comprehensive pan-organ transplantation transcriptomic atlas, was used to decode the transcriptional hallmarks of allograft outcomes(23). The ENCODE encyclopedia was used to decode functional gene regulatory elements(24).

### Single-cell RNA sequencing from lung explants

Lung explants were retrieved at the time of transplantation and immediately transferred to the laboratory in Perfadex medium (XVIVO) on ice for cellular dissociation. In total, 4 lungs from different participants were analyzed: 2 bronchiolitis obliterans syndrome (BOS) and 2 healthy donor lungs (HC). Single-cell RNA-sequencing (scRNAseq) was performed from these using the Chromium 10X GEM-X library and a Nova-Seq 6000 sequencing system – the sample preparation protocol and scRNAseq analytical details are available in the Supplementary methods.

## Results

### Demographics and clinical characteristics

From the 392 pairs included in the GenCOLT biocollection, 387 pairs (99%) passed the DNA and GWAS quality controls (Fig.1). We restricted our analyses to European individuals who developed CLAD (n=67), or with stable lung function after 5 years of follow-up (non-CLAD control group, n=124). Demographic and clinical characteristics of the recipients and donors were overall very similar between the CLAD and non-CLAD subgroups, except for older age and shorter follow-up time observed in CLAD patients (Tables 1 and 2, respectively). Additional details are available in the Supplementary results.

**Table 1:**
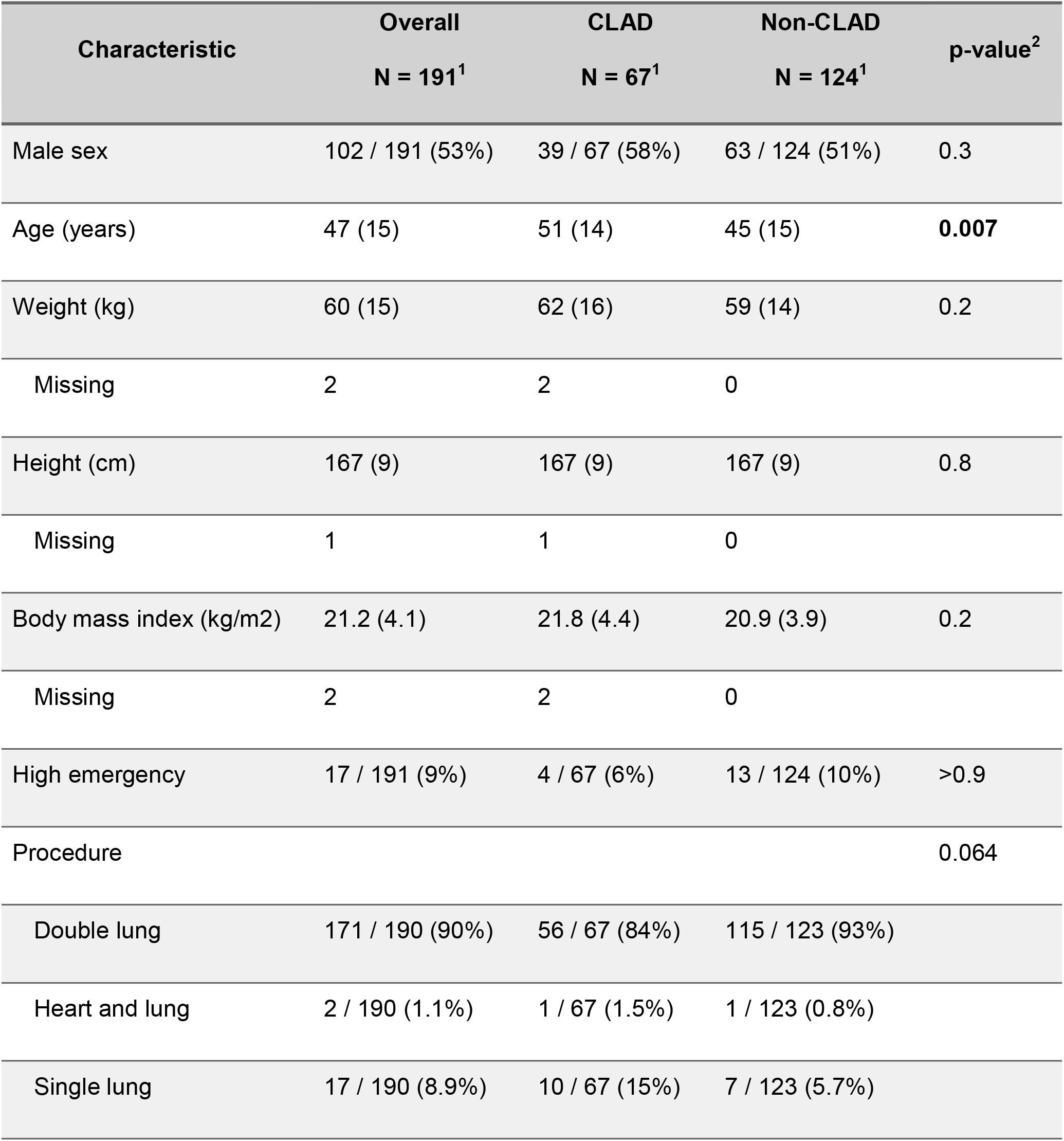
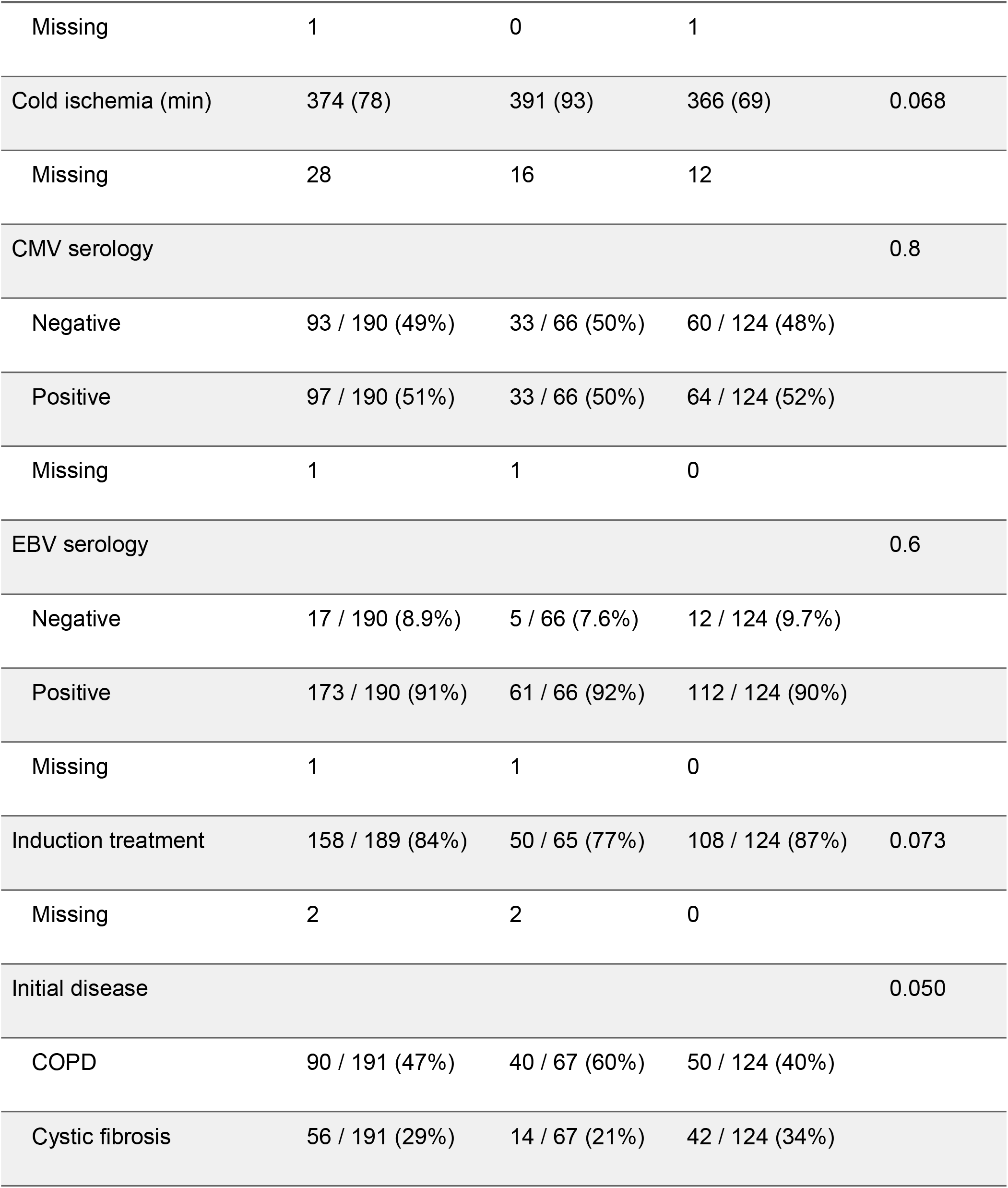
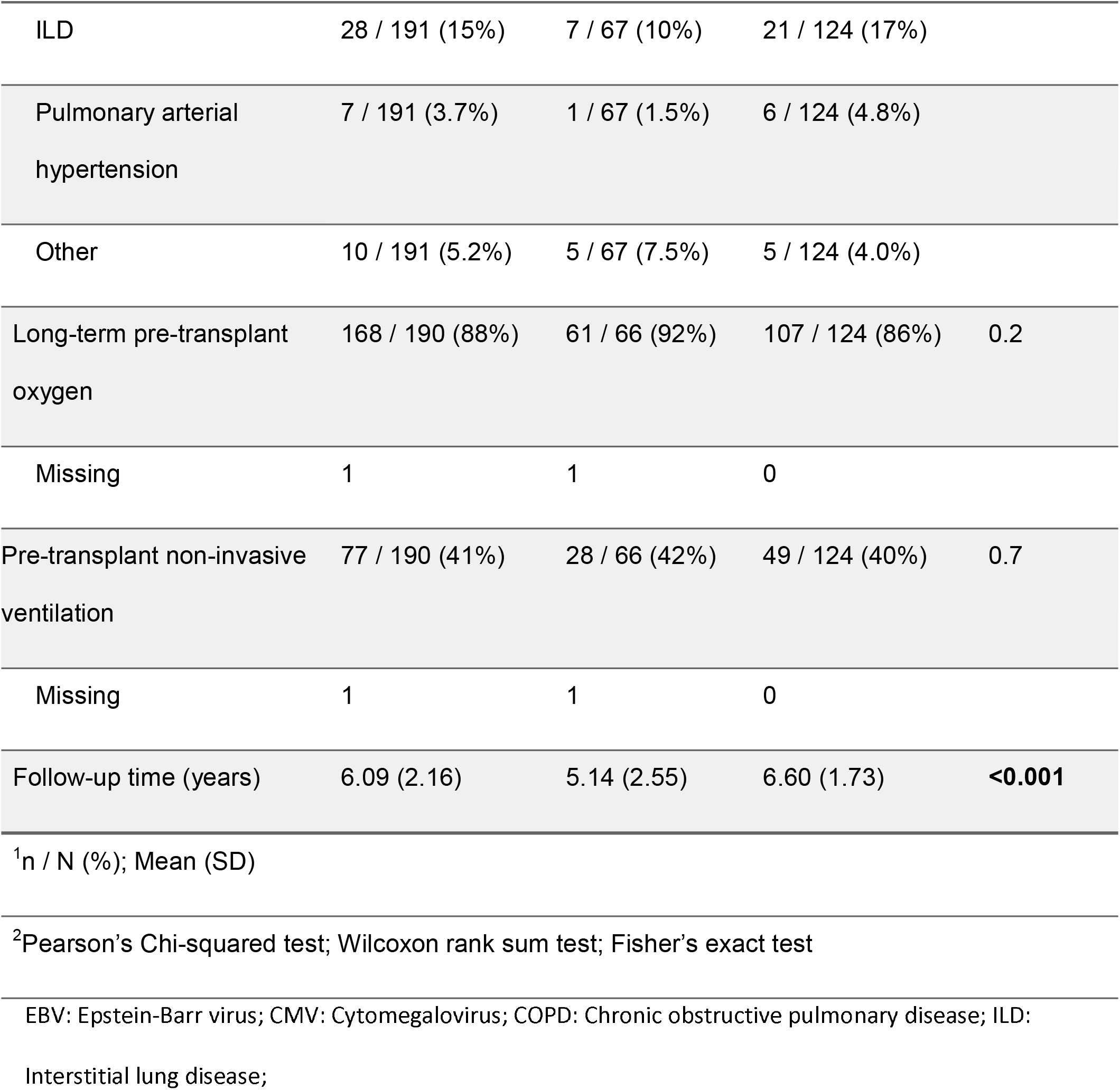
Demographic table for the recipients. Overall, all characteristics were similar between the CLAD and non-CLAD subgroups, except for older age and shorter follow-up time observed in patients experiencing CLAD.

**Table 2:**
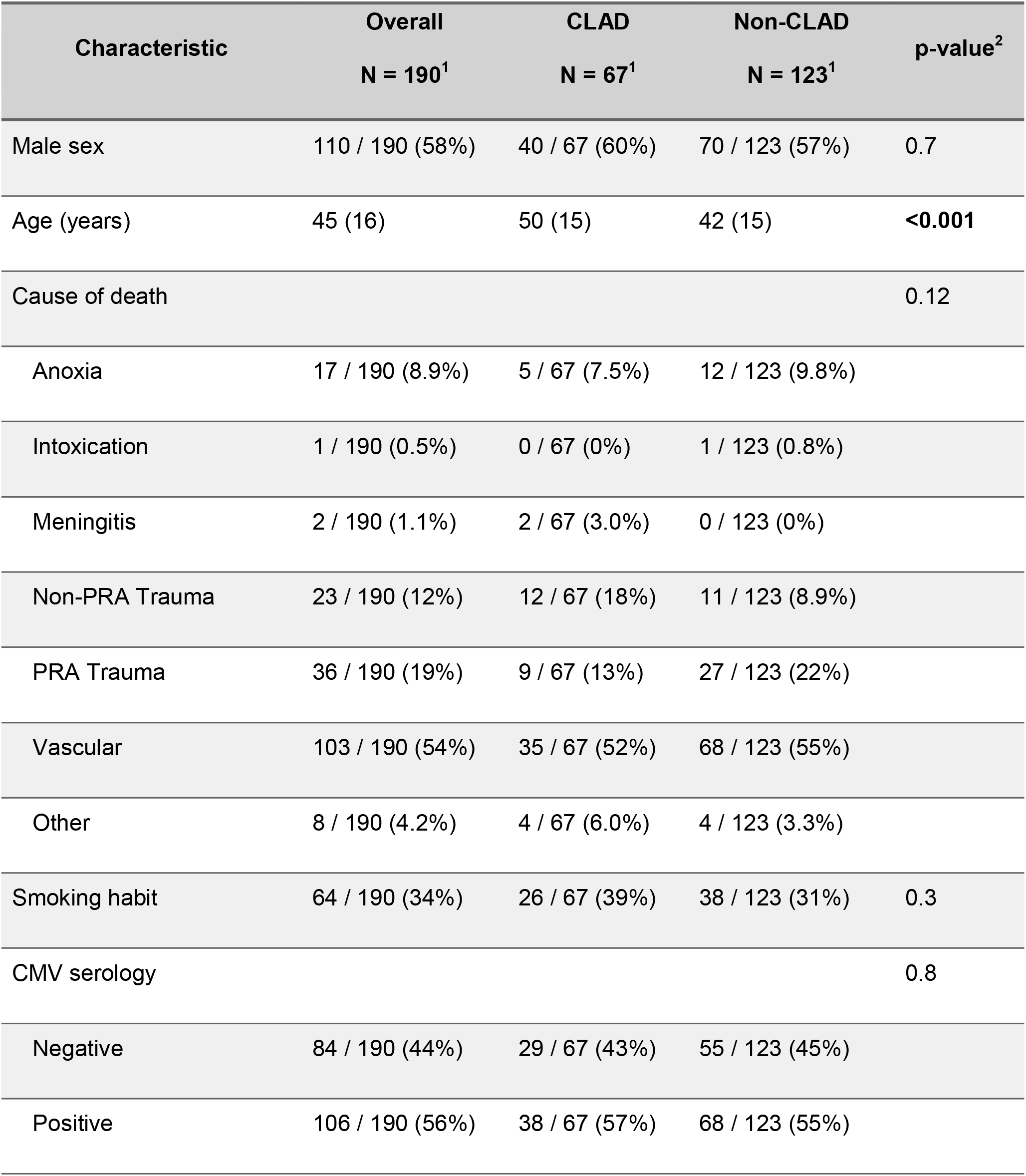
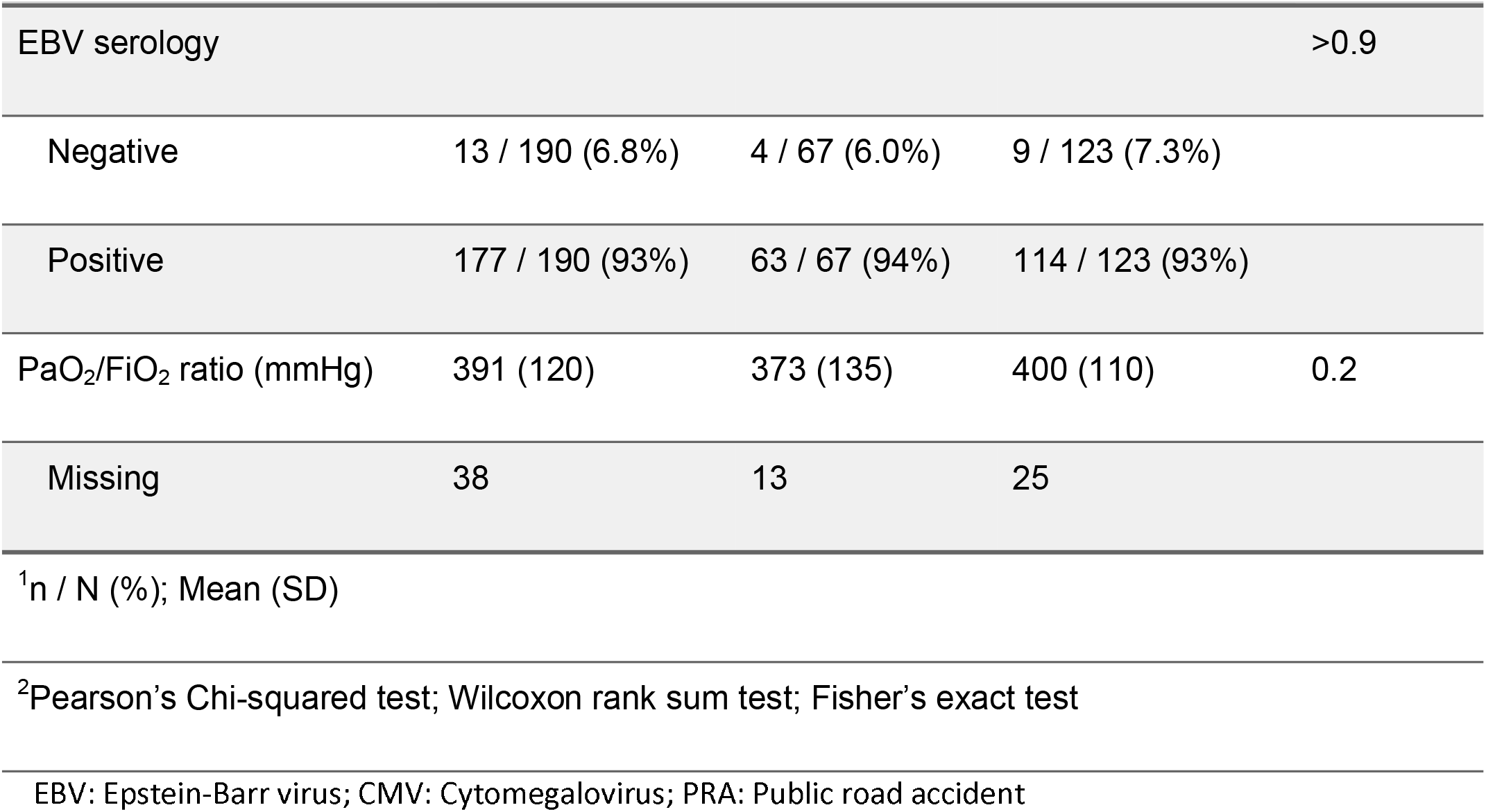
Demographic table for the donors. Overall, all characteristics were similar between the CLAD and non-CLAD subgroups, except for older donor age for patients experiencing CLAD.

### HLA-centric association analysis

We first ran an association analysis with CLAD for each *HLA* allele from each major classical *HLA* gene at three levels: recipient-only *HLA* alleles, donor-only *HLA* alleles, and donor-recipient *HLA* allele mismatches (Fig.S2A-C), and did not observe any significant association. We then examined donor-recipient allelic mismatches at each individual *HLA* loci and identified a significant association with CLAD risk for increasing *HLA-A* mismatches (p=0.01, β=0.87, Fig.S3). Finally, we analyzed total allelic and total epitopic *HLA* mismatch scores across all class I, all class II, or all class I+II *HLA* genes. No significant associations were observed for total allelic mismatch scores, but a significant association with CLAD emerged for the increasing load of total HLA epitopic mismatches (p=0.026, β=0.03), which was driven by the class I epitopic mismatch score (p=0.004, β=0.06, Table S1).

### GWAS revealed 2 significant associations within the recipients’ genome with CLAD

After GWAS quality controls and SNP imputation, 4.5 million SNPs with MAF≥10% were tested for association with CLAD (statistical power >75% for OR≥2.5, Fig.S1), considering recipient-only genotypes, donor-only genotypes, and donor-recipient mismatches. The QQplots revealed an absence of systemic deviation from the null hypothesis for the recipients’ genotypes analysis (Fig.S4A), and a limited power for the donors’ genotypes and the donor-recipient mismatch analyses (Fig.S4B-C). In this context, no significant associations were identified when investigating the donors’ genotypes or the donor-recipient mismatches (Fig.2). However, the analyses of the recipients’ genome revealed 65 SNPs from 2 independent chromosome 10 loci significantly associated with CLAD (FDR q-value <5%; Fig.2 and Table S2): the top rs10734030 SNP (FDR=0.017; p=5.05×10^-7^; 38.5% in CLAD vs. 66.3% in controls; OR=0.35) located in a *PLXDC2* intron (Fig.3A-B), and the second locus with the top rs4526713 SNP (FDR=0.017; p=9.86×10^-7^; 65.9% in CLAD vs. 38.2% in controls; OR=2.83) located in a *ZNF518A* intronic region and downstream *BLNK* (Fig.4A-B, Table S2).

**Figure 2:**
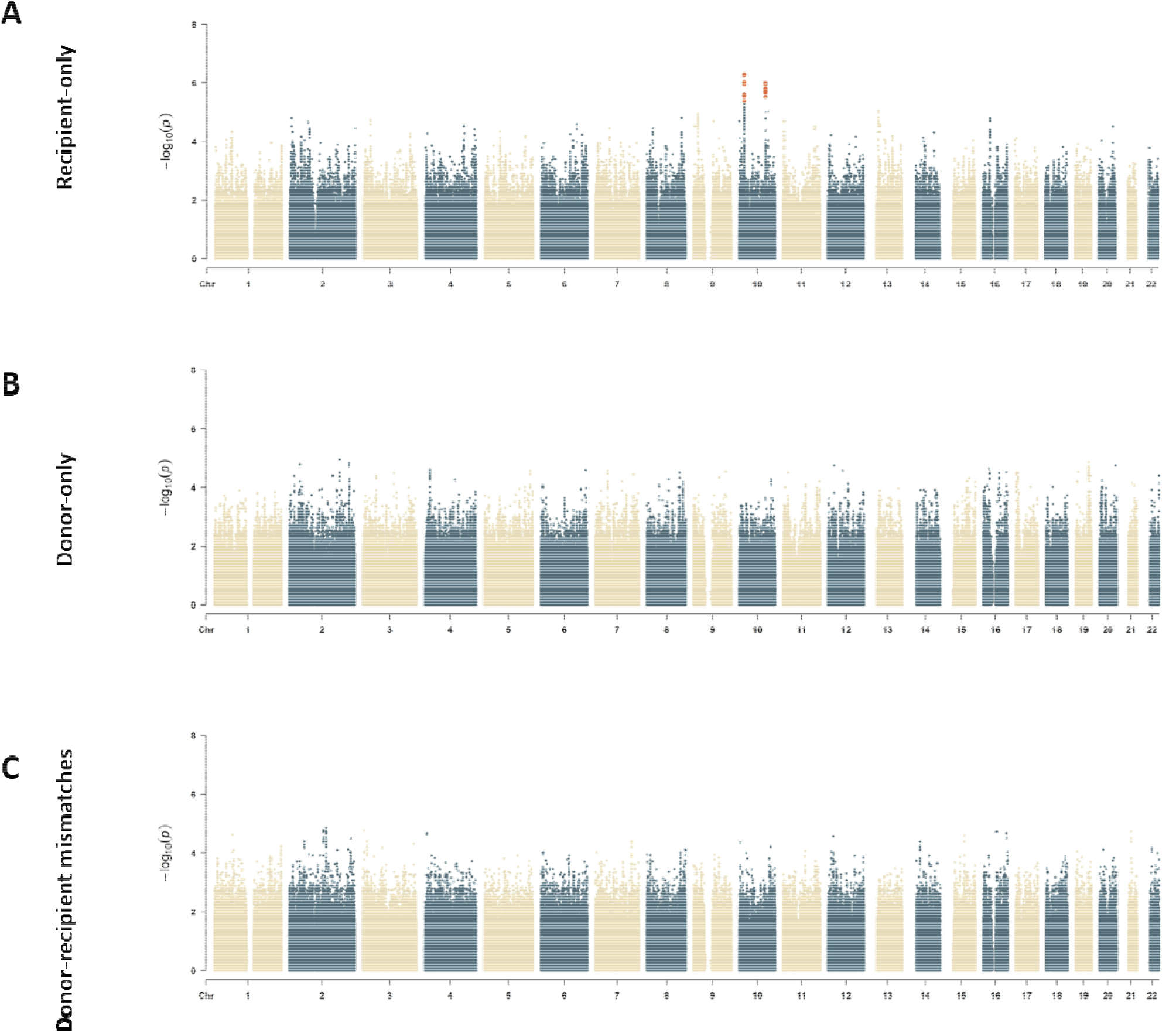
Manhattan plots summarizing the genome-wide association (GWAS) results with chronic lung allograft dysfunction (CLAD). Each dot represents one of the 7.3 million SNPs tested for association. The x-axis shows the SNP position along the genome, and the y-axis shows the statistical significance as -log_10_ of P-values. The SNPs passing the FDR<5% significance threshold are highlighted in bold and red.

**Figure 3:**
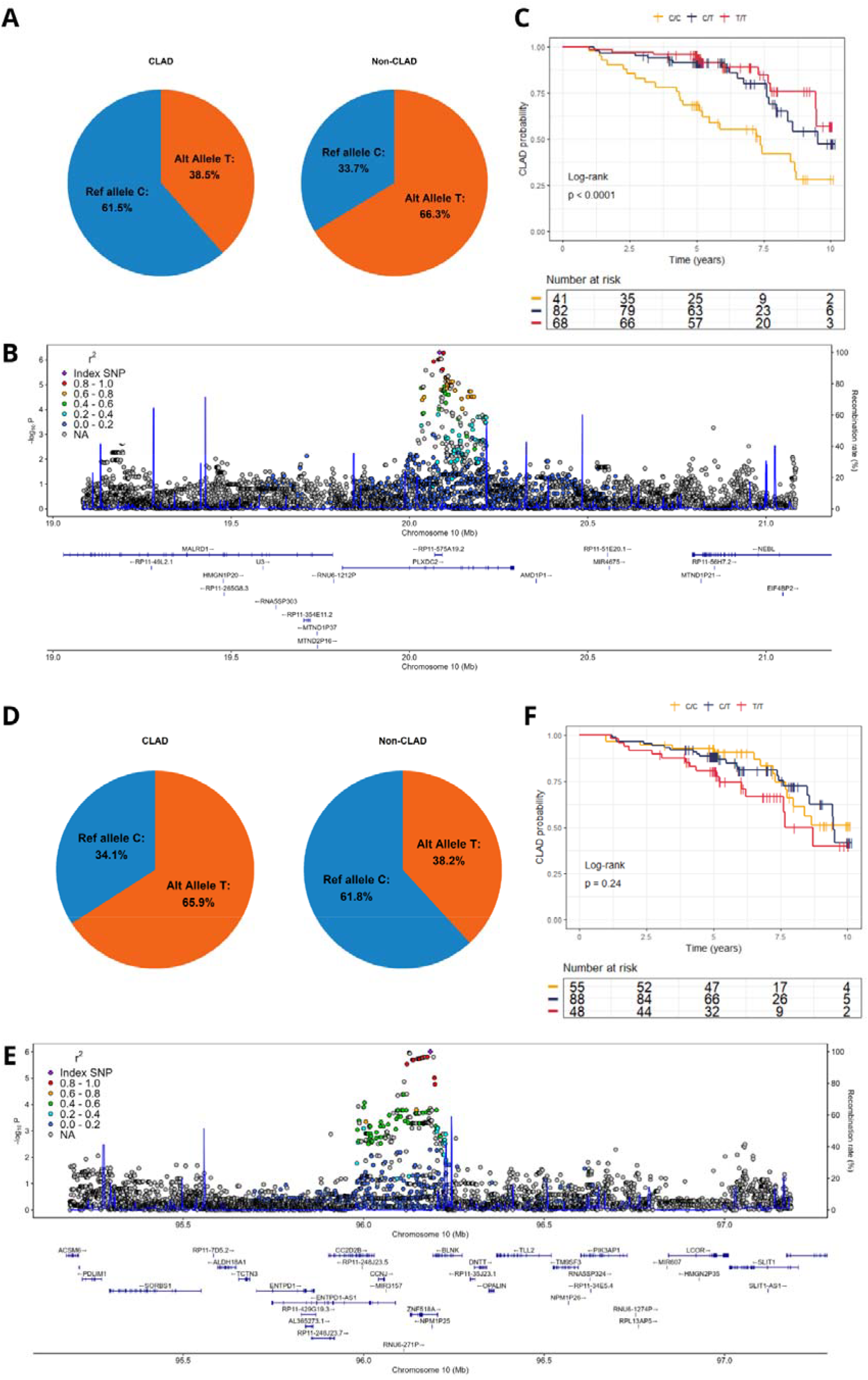
Comprehensive examination of top significant signals. A: Cases vs controls frequencies of the top SNP in the *PLXDC2* locus. B: Survival analysis of the top SNP in the PLXDC2 locus using a Kaplan-Meier estimator. C: Regional association plot of the *PLXCD2* locus. D: Cases vs controls frequencies of the top SNP in the *ZNF518A* locus. E: Survival analysis of the top SNP in the *ZNF518A* locus using a Kaplan-Meier. F: Regional association plot in the *ZNF518A* locus.

**Figure 4:**
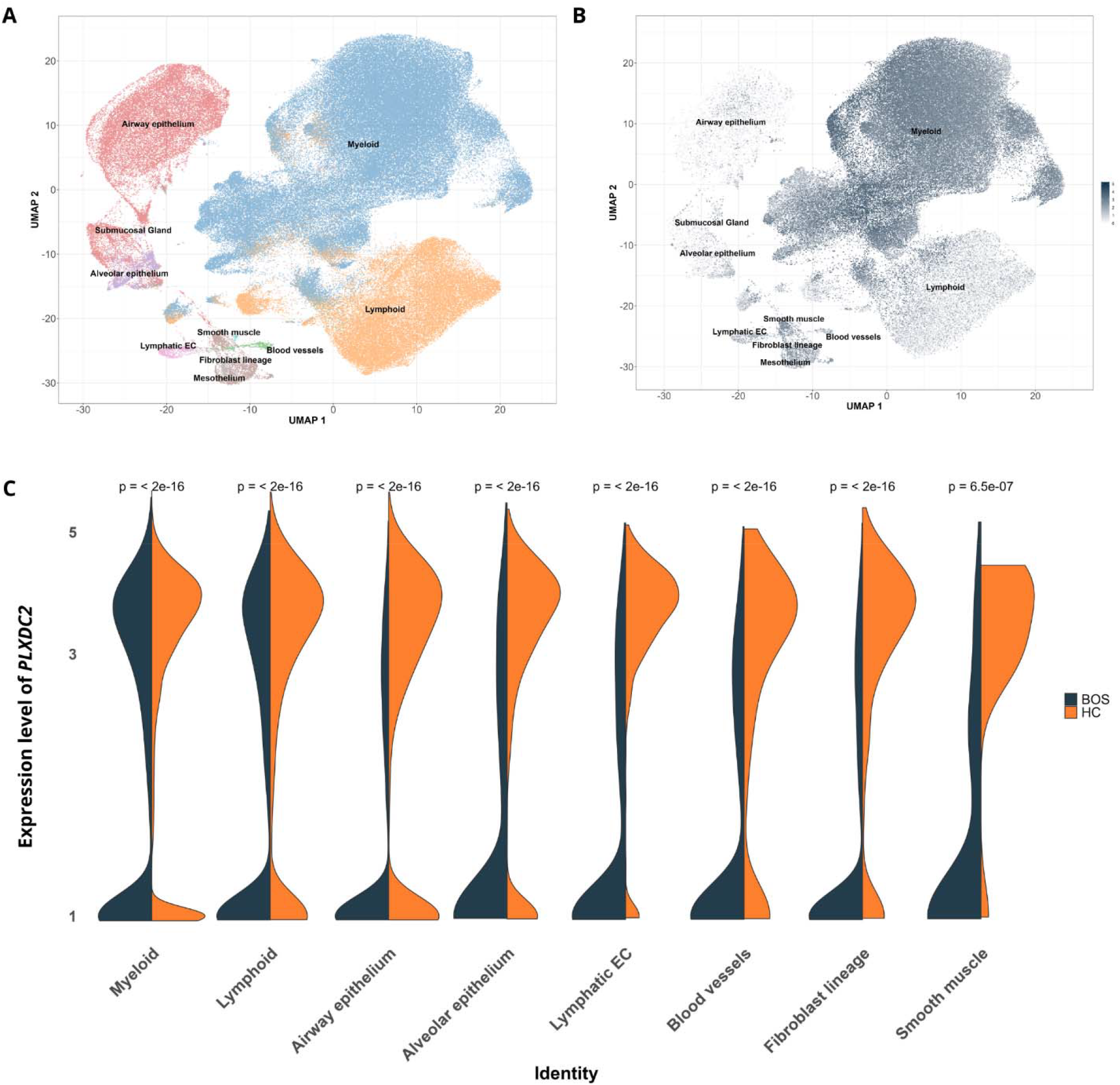
Single-Cell profiling of *PLXDC2* in pulmonary explants. A: Two-dimension reduction map (UMAP) of the single-cell transcriptomics datasets highlighting the main lung cell populations in BOS and healthy samples. B: UMAP overlaid with expression of *PLXDC2* in the single-cell transcriptomics dataset. C: Differential *PLXDC2* expression in lung explants from 2 BOS patients vs. 2 healthy control (HC). There are too few cells for submucosal gland (n=1 cells) and mesothelium (n=8 cells) which is why they are not represented in the violin plot. EC: Endothelial cells.

To consider time of CLAD occurrence, we performed a survival analysis using Kaplan-Meier plots and Cox models for the two top SNPs from each independent chromosome 10 locus (Fig.3C-4C). The rs10734030 *PLXDC2* SNP was significantly associated with time-to-CLAD (HR=0.48 [0.33-0.70]; p≤0.001), with patients carrying the T allele (TT or CT genotypes) exhibiting significantly longer protection from CLAD compared to those carrying the CC genotype. In contrast, the rs4526713 *ZNF518A/BLNK* SNP was not associated with time-to-CLAD in a survival analysis framework (HR=1.31 [0.90-1.91]; p=0.2).

Interestingly, we observed a different frequency distribution for the rs10734030-C allele across continents (from 0-3% in East Asia and Africa, up to >35% in Europe), which could indicate local adaptation or evolutionary selective pressure after the out-of-Africa exodus according to our selection signature analysis (Fig.S5-S6 and Supplementary results).

To circumvent the limited statistical power in our donor-recipient mismatch analysis, we summarized the overall mismatch burden across the genome or in relevant lung genes into non-HLA mismatch scores, but none of these polygenic scores were associated with CLAD (Table S3, see Supplementary for more details).

### Functional exploration of the top signals

Both the *PLXDC2* and the *ZNF518A/BLNK* SNPs were located in non-coding and putative regulatory regions. Both top independent SNPs were described as eQTLs (*i*.*e*. genetic variants associated with differential gene expression levels) in cell types relevant for CLAD pathogenesis. rs10734030 was described as an eQTL for *PLXDC2* in different tissues, including pancreas (p=1.02×10^-9^, with the T allele associated with a higher *PLXDC2* expression). Interestingly, the entire chromosome 10 *PLXDC2* region is enriched for regulatory elements (Fig.S7). Similarly, the rs4526713 SNP was reported as an eQTL for *ZNF518A* in multiple tissues, including lung (p=4.32×10^-7^, with the T allele associated with an increased *ZNF518A* expression) and lymphocytes (p=1.81×10^-6^, with the T allele associated with a lower *ZNF518A* expression), and as an eQTL for *BLNK* in lymphocytes (p=1.03×10^-6^, with the T allele associated with a lower *BLNK* expression).

Using scRNAseq from two explanted lungs with BOS and two healthy lungs, we highlighted the main lung and immune cell types (Fig.5A). The *PLXDC2* gene was predominantly expressed in myeloid cells (n=80,309 cells), airway epithelium (n=14,453 cells) and lymphoid cells (n=11,478 cells, Fig.5B). When comparing *PLXDC2* cell-specific expression between patients experiencing BOS vs. healthy controls, *PLXDC2* was systematically significantly down-regulated in BOS (p≤2.0×10^-16^, Fig.5C). This is consistent with rs4526713-T being associated with higher *PLXDC2* expression and showing a protective effect from CLAD. *ZNF518A* and *BLNK* were not expressed in our lung explant dataset. However, a differential expression of both genes was reported in lungs during allograft rejection in an independent meta-analysis (Fig.S8), reinforcing the fact that the rs4526713 association with CLAD risk could be mediated through gene expression regulation.

## Discussion

In this report, we systematically explored common genetic variants for association with CLAD in a unique cohort of 392 donor-recipient lung transplant pairs. First, we identified a significant association with CLAD for the *HLA-A* locus mismatches, with an increased risk of 1.2 for each allelic difference. This deleterious effect was also captured by the HLA class I and the total HLA epitopic mismatch scores. Our results further complement the literature on *HLA* mismatch associations with LT outcomes. To our knowledge, it is the first time that significant associations of *HLA-A* locus and epitopic mismatches are reported with CLAD while finely accounting for genetic ancestry, as it has not been considered in previous reports(9).

We then investigated the association of recipients, donors, and donor-recipient mismatch SNPs with CLAD and identified 65 recipients’ SNPs from two independent loci associated with CLAD. First, the *PLXDC2* signal appears protective from CLAD (rs10734030-T, 38.5% CLAD vs. 66.3% in controls), with a sustained effect over post-transplantation follow-up time (T carriers exhibited a better CLAD-free survival). The second signal conferred a 2.8 increased risk for developing CLAD (rs4526713-T, 65.9% in CLAD vs 38.2% in controls) without an additional effect over time under a survival analysis framework.

When exploring the recent selective signatures within different ancestral groups, a positive selection was identified for the *PLXDC2* SNP within Europeans. A positive selection signal implies that the allele may have been favored during recent evolution due to possible beneficial effects. This evolutionary footprint often indicates functional relevance, and in this case, it could point to a key role in immune-related processes, reflecting past adaptation to pathogenic pressures.

Interestingly, the *PLXDC2* genetic variants have previously been associated with bronchopulmonary dysplasia(25). It was also reported that PLXDC2 elevates phosphorylated cortactin (p-Cortactin) levels in tissues by interacting with the protein tyrosine phosphatase 1B (PTP1B) thereby inhibiting its ability to dephosphorylate p-Cortactin(26). Cortactin is a central regulator of actin cytoskeleton dynamics and barrier integrity, and cortactin phosphorylation has been implicated in multiple pulmonary diseases, including acute lung injury, asthma, COPD, bronchopulmonary dysplasia, and aspirin-exacerbated respiratory disease(27). Recently, a GWAS identified polymorphisms in the cortactin gene associated with the FEV_1_/FVC ratio and alveolar epithelium modelling(28). In this relevant functional context, our in-silico explorations suggested that the rs10734030 SNP could impact the *PLXDC2* gene expression levels, and we reported a consistent decreased *PLXDC2* expression within all lung cell types during BOS, including monocytes where *PLXDC2* is primarily expressed. We can hypothesize that the rs10734030 SNP association with CLAD could be driven by the dysregulation of *PLXDC2* with a subsequent impact on cortactin phosphorylation and tissue injury. Further validation in an independent cohort and in a larger number of lung explants is needed to confirm these observations.

The second independent locus lies within the *ZNF518A* gene, and the lead SNP was associated with its gene expression. ZNF518A protein acts as a key regulator in the formation of pericentromeric heterochromatin, particularly influencing α-satellite DNA regions, which is crucial for maintaining genomic stability and proper chromatin organization(29). While there is no direct evidence linking ZNF518A to CLAD physiopathology, modifications of *ZNF518A* expression could hypothetically impact chromatin remodeling, which may influence the risk of rejection. Notably, the top rs4526713 SNP serves as an eQTL for *ZNF518A* in immune cell populations, highlighting a potential mechanism whereby the ZNF family may critically shape immune response regulation(30). Importantly, the lead SNP within this locus was also associated with *BLNK* expression, which plays a critical role in B cell development. *BLNK* gene expression was previously associated with lung cancer(31) and severe Covid-19(32), underlying the relevance of this locus for lung tissue injury. Importantly, *BLNK* expression has also been associated with chronic graft-versus-host disease(33) and with operational tolerance and graft rejection in kidney transplantation(36,37,38), highlighting its potential key role in immune response regulation. Both chromatin remodeling and B cell regulation could be important for lung allograft rejection risk, and our genetic association and *in-silico* explorations alone cannot dichotomize the causal underlying gene(s), hence calling for further functional investigations.

Akin to what was proposed in kidney transplantation(35), we summarized the overall burden of potential immunogenic antigens presented by the donor as non-self to the recipient into non-HLA mismatch scores, but none of the tested SNP subset configurations were associated with CLAD risk in our cohort (Fig.S9). This important result calls for considering novel biological hypotheses (including the role of regulatory and rare variants), innovative SNP selection tools (*e*.*g*. AI-based), and for recruiting larger donor-recipient cohorts in LT.

The main limitation of the present study was its small sample size, despite being the largest to date on lung transplant genetics. A power analysis indeed revealed that our cohort exhibited ≥75% statistical power for discovery of SNPs associated with CLAD with a MAF≥10% and an effect size ≥2.5 (Fig.S1). Both our significant signals are indeed within this range. A larger sample size or an independent cohort will be necessary to confirm these signals and potentially reveal additional loci. Functional investigations would also be warranted to pinpoint the causative molecular mechanisms underlying CLAD for each highlighted locus. In addition, GenCOLT primarily consists of individuals of European descent, limiting our ability to draw conclusions about genetic risk factors for lung graft rejection beyond the European population. To overcome these limitations, we are actively involved in the International Genetics and Translational Research in Transplantation Network (iGeneTRAiN)(36,37), which is the largest global genetic initiative dedicated to understanding transplant rejection and related complications with organ-specific and cross-organ strategies.

To conclude, our genomic association study of CLAD in European lung transplanted patients has revealed two novel independent significant loci from the recipients. Together, these findings offer deeper insights into the genetics foundations underlying CLAD, paving the way for molecular and cellular characterization of its pathophysiology. They also underscore the need for larger-scale genomic studies that integrate detailed clinical outcomes to drive progress toward targeted interventions.

## Abbreviations

ARAD: azithromycin responsive allograft dysfunction
BMI: body mass index
BOS: bronchiolitis obliterans syndrome
CLAD: chronic lung allograft dysfunction
COLT: cohort in lung transplantation
COPD: chronic obstructive pulmonary disease
DNA: deoxyribonucleic acid
FEV1: forced expiratory volume in 1 second
GWAS: genome-wide association study
HLA: human leukocyte antigen
iHS: integrated haplotype homozygosity score
LT: lung transplantation
MAF: minor allele frequency
PCA: principal component analysis
PMRA: precision medicine research array
QC: quality control
RAS: restrictive allograft syndrome
RNA: ribonucleic acid
SNP: single nucleotide polymorphism

## Acknowledgments

The Cohort in Lung Transplantation was funded by *Vaincre La Mucoviscidose, l’Association Grégory Lemarchal*, the French Research Ministry (*Agence Nationale de la Recherche* grant), the French Government (*Programme Hospitalier de Recherche Clinique* – DGOS 20-11), the European Union (FP7 collaborative project HEALTH.2012.2.1.2-1 – grant agreement number: 305457), the *Institut de recherche en santé respiratoire des Pays de Loire* and Nantes Metropole. The GenCOLT study was funded by the Nantes university hospital. PA Gourraud initated this genetic collaboration through the Inserm ATIP-Avenir Fellowship and Connect-Talents Award (Region Pays de La Loire, France). The study sponsor(s) or funder(s) had no role in the study design, in the collection, analysis, and interpretation of data, in the report writing and, in the decision, to submit the article for publication. Researchers were independent from funders and all authors, external and internal, had full access to all data (including statistical reports and tables) in the study and can take responsibility for the integrity of the data and the accuracy of the data analysis. S. Brocard benefited from the support of IMT Atlantique, Centrale Nantes and Institut de Recherche en Santé Respiratoire des Pays de la Loire for his PhD research. We thank the participating patients and their families, whose trust, support, and cooperation were essential for the collection of the data used in this study. We thank the COLT Consortium for their organizational achievement and the LUNG O_2_ cluster. The authors thank everyone who helped in the design, collection, genotyping experiments, data cleaning and analyses. We are grateful to the Genomics Core Facility GenoA, member of Biogenouest and France Genomique (ANR-11-INBS0013), for the use of its resources and technical support. Finally, we thank the biological resource center for biobanking (CHU Nantes, Hôtel Dieu, *Centre des ressources biologiques* (CRB, BRIF: BB-0033-00040)).

## Data availability statement

The GenCOLT data are available from our CR2TI team. Data and summary statistics are available upon request following approval from the GenCOLT steering committee and ethics committee to ensure data protection and privacy in compliance with French and European laws. Collaborations are encouraged through specific research projects using GenCOLT data or through enriching the existing cohort with new patients. The GWAS summary statistics were deposited to the GWAS Catalog (GCP001413). Potential collaborators are invited to contact the primary investigator Sophie Limou.

## Author contributions

Initial study design, S.L. and A.T.; Providing DNA samples and patients monitoring, B.R.P., B.C., X.D., L.F., J.L.P., A.R., T.V., C.K., C.M., M.S., N.C., A.M., D.L. and A.T.; Data curation, genomic and statistical analyses, S.B.; Methodology and software, S.L., M.M., V.M., N.B.S., A.D. and N.V.; HLA expertise, N.V., N.B.S., and P.A.G; scRNAseq data and analysis, Le.B., P.H. and La.B.; Writing original draft, S.B.; Manuscript review and editing, S.B., S.L., A.T. and M.S. All authors have read and agreed to the published version of the manuscript.

