## Supplemental for "Genome-wide association study reveals two novel genetic loci associated with chronic lung allograft dysfunction"

by Brocard *et al.*

### Figures

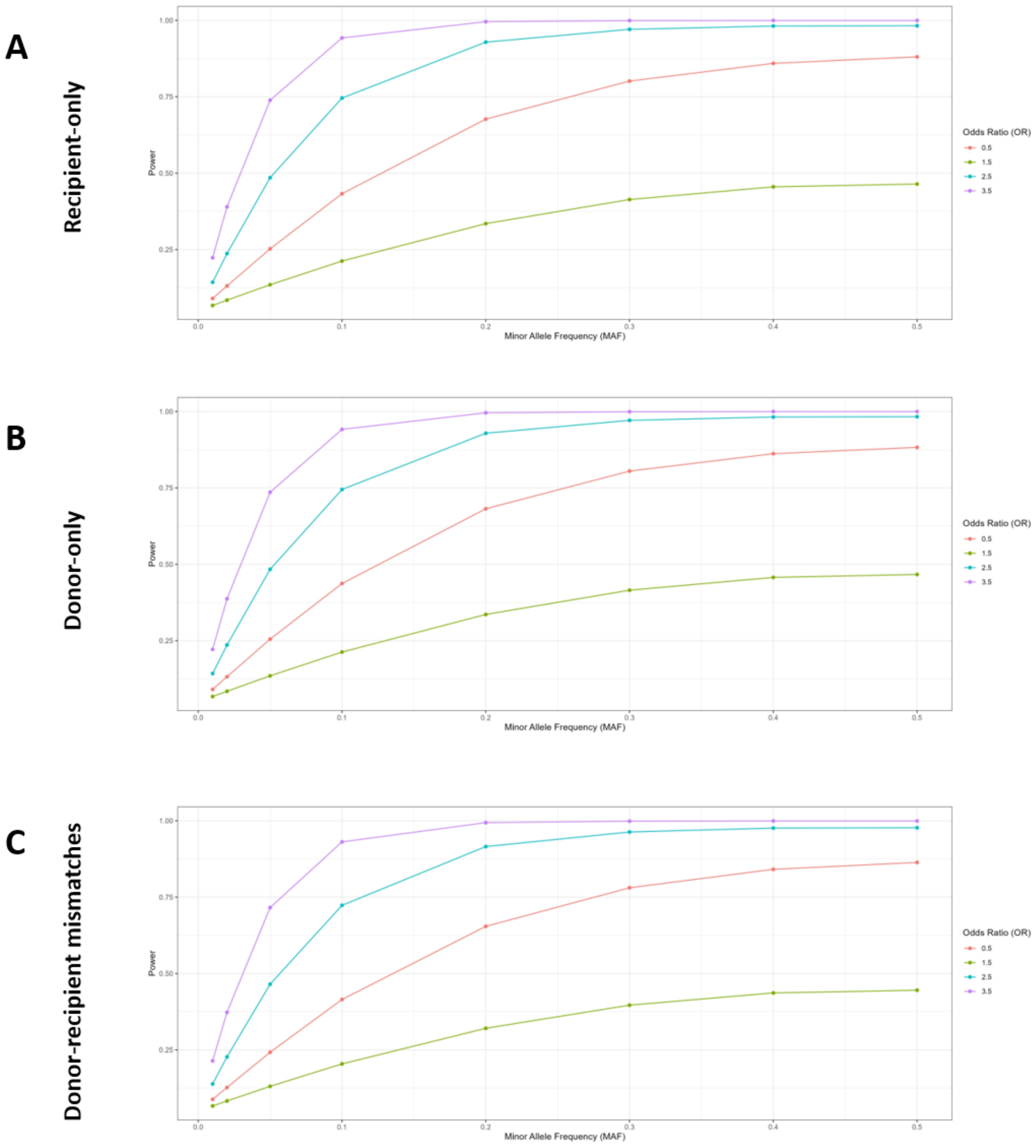


*Figure S1: Power analyses of our GWAS study design*

The statistical power of our study was evaluated according to different hypotheses of odds-ratio and minor allele frequency (MAF) using the genpwr package (v1.0.4)(1) for recipients-only (**A**), donors-only (**B**) and donor-recipient mismatches (**C**) association analyses. Due to the limited statistical power of our cohort, we restricted our GWAS analyses to SNPs with a MAF≥10% to increase the likelihood of detecting true positive associations: power >75% for OR≥2.5 and >90% for OR≥3.5.

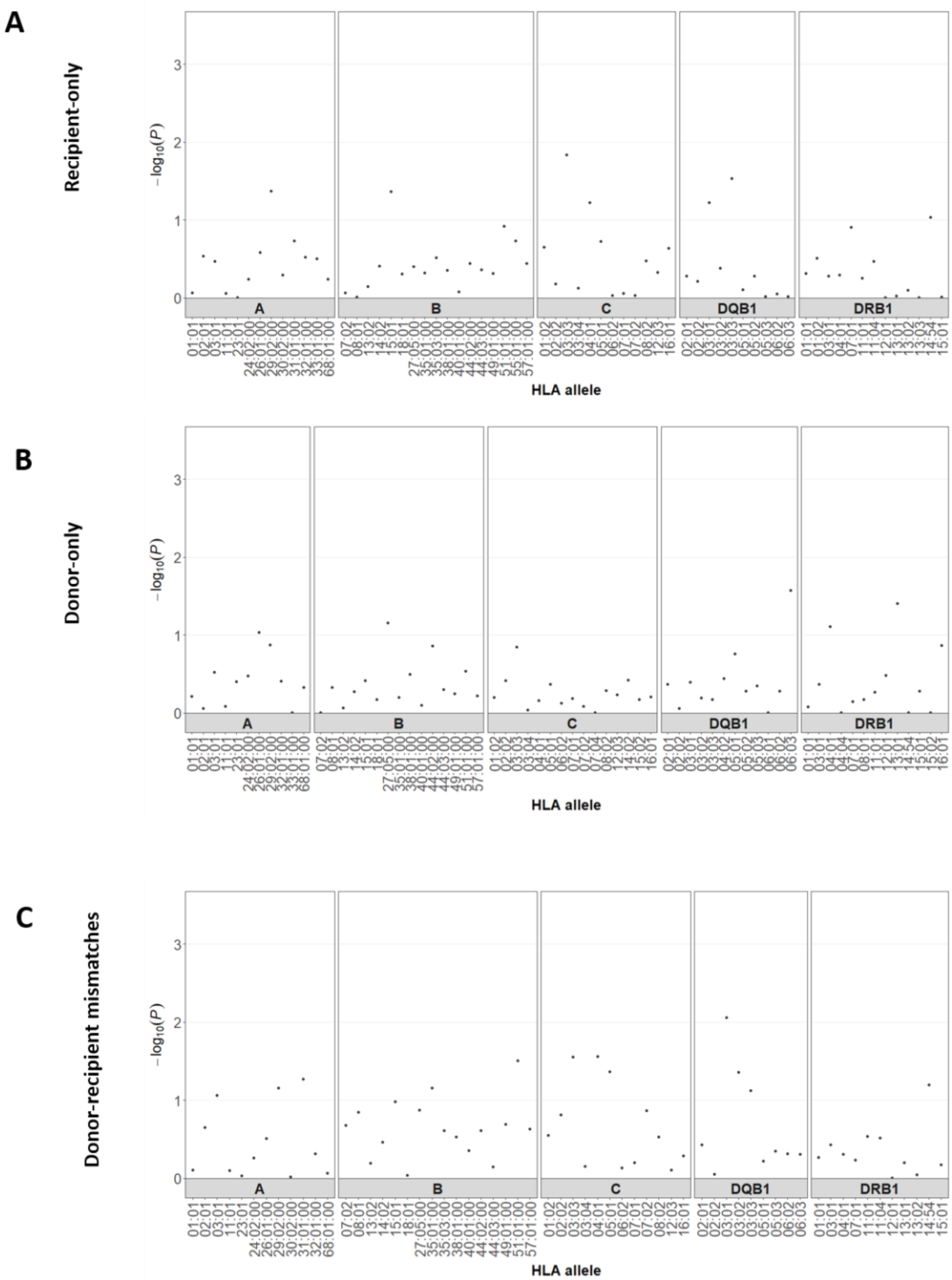


*Figure S2: HLA-centric association analysis with CLAD*

Manhattan plots for *HLA* allele association study with CLAD for the 5 major classical *HLA* genes (*HLA-A, HLA-B, HLA-C, HLA-DRB1* and *HLA-DQB1*) focusing on recipients-only (**A**), donors-only (**B**), and donor-recipient mismatches (**C**). Each dot represents an *HLA* allele for one of the 5 major classical *HLA* genes with its name reported on the x-axis, when the y-axis shows the statistical significance as $-\log_{10}$ of the P-value. The Bonferroni significant threshold was set at $p=10^{-4}$ (beside the graph limits). No *HLA* allele from the recipients, donors, or donor-recipient mismatches was significantly associated with CLAD in our cohort.

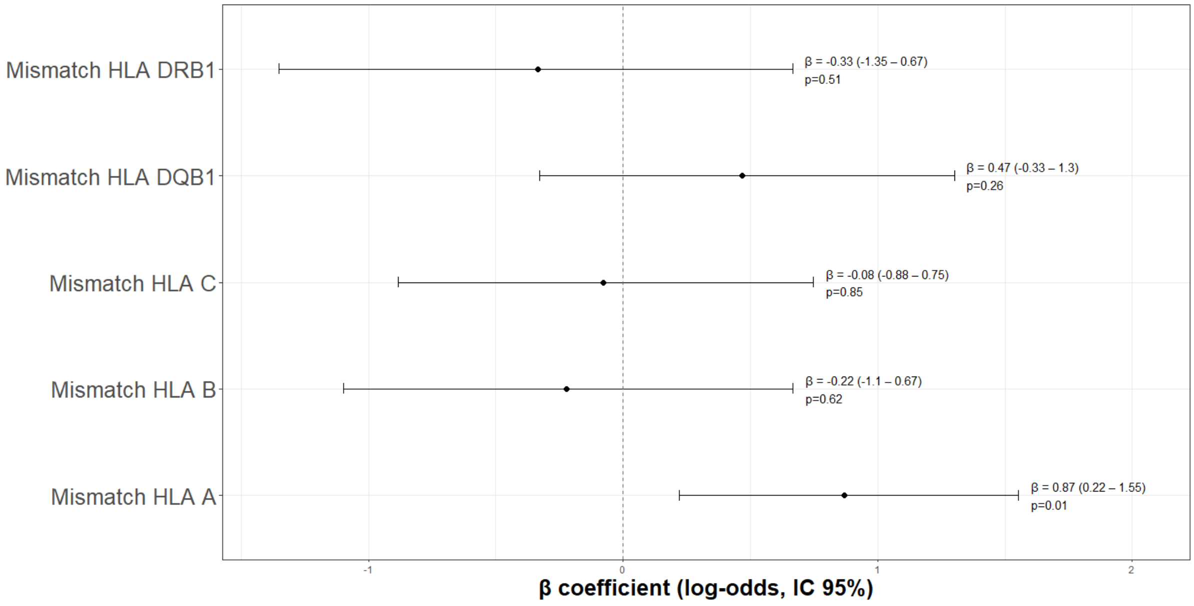


*Figure S3: Donor-recipient allelic mismatches association with CLAD*

Forest plot of the association between classical *HLA* locus (*HLA-A, HLA-B, HLA-C, HLA-DQB1* and *HLA-DRB1*) allelic mismatch (0/1/2) and CLAD, corrected for age, sex, genetic ancestry (five first principal components), and initial disease. We observed a significant association for *HLA*-A mismatch with CLAD risk (p=0.012).

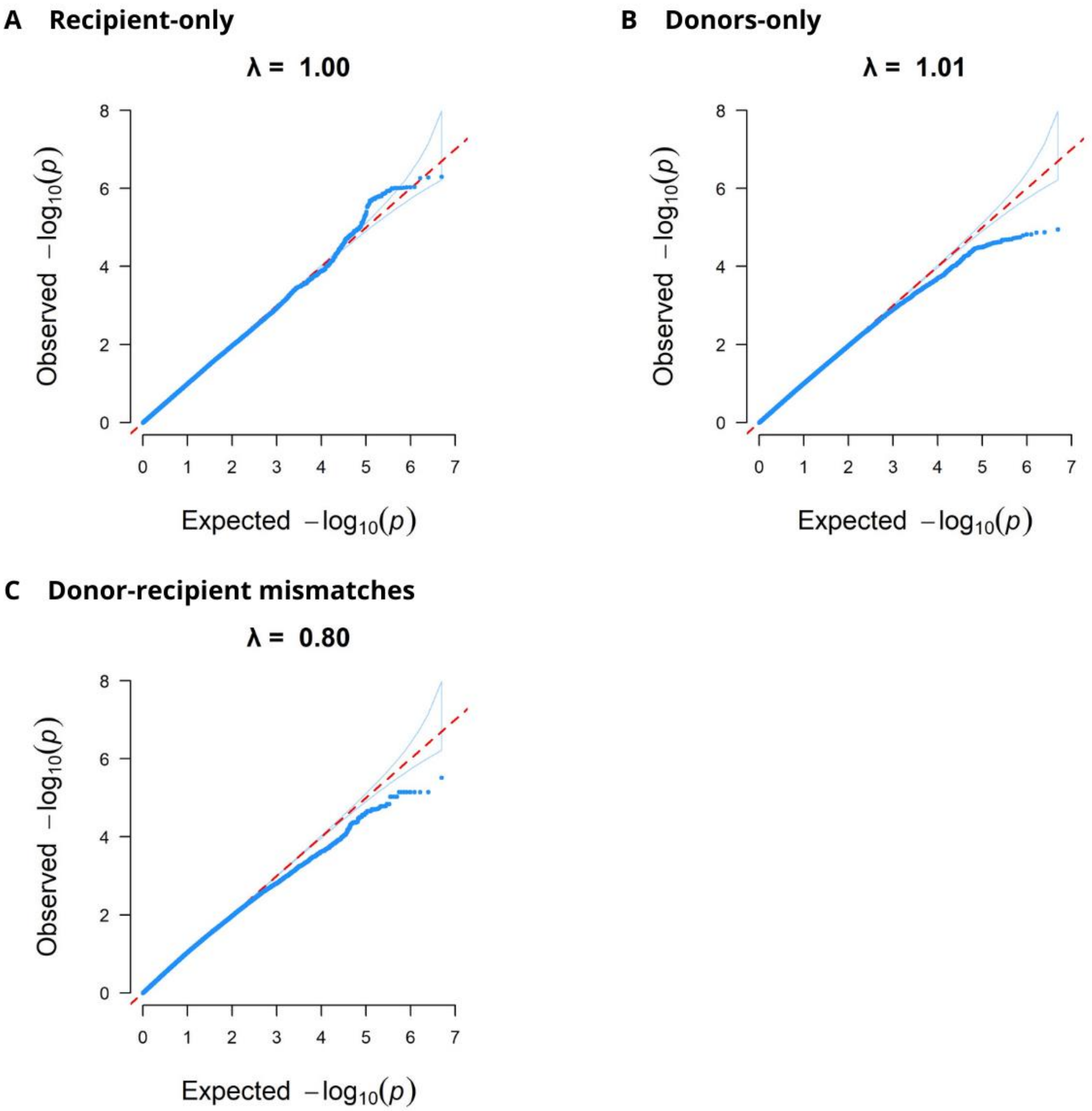


*Figure S4: Quantile-Quantile plots summarizing the GWAS results with CLAD.*

QQplots corresponding to the Manhattan plots presented in Figure 2. The null distribution is shown as a red dotted line. The donors and donor-recipient mismatches analyses appear slightly underpowered.

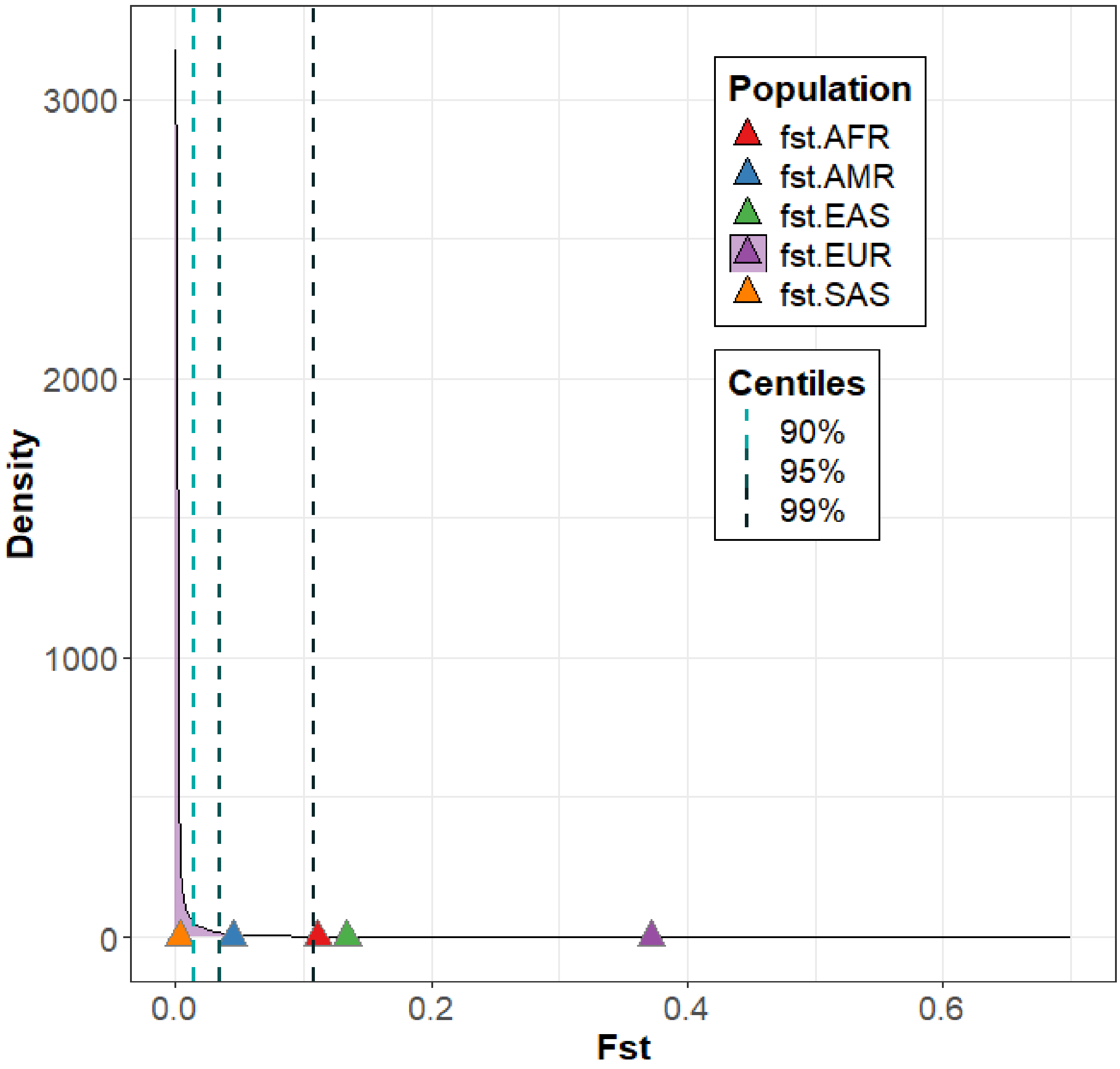


*Figure S5: Top CLAD-associated SNP's $F_{st}$ values within the $F_{st}$ index genome distribution*

The fixation index statistics ($F_{ST}$) was calculated for each SNP within the human genome between each ancestral continental population vs. the others, and extracted from the 3DSNP database(2). The $F_{ST}$ values for our top rs10734030 SNP associated with CLAD in the recipient GWAS are depicted with colored triangles. Interestingly, a strong positive selection signature was observed for the *PLXDC2* SNP in Europeans (frequency >35% vs. 0-3% in East Asians and Africans).

*AFR: African; AMR: American; EAS: East Asian; EUR: European; SAS: South Asian.*

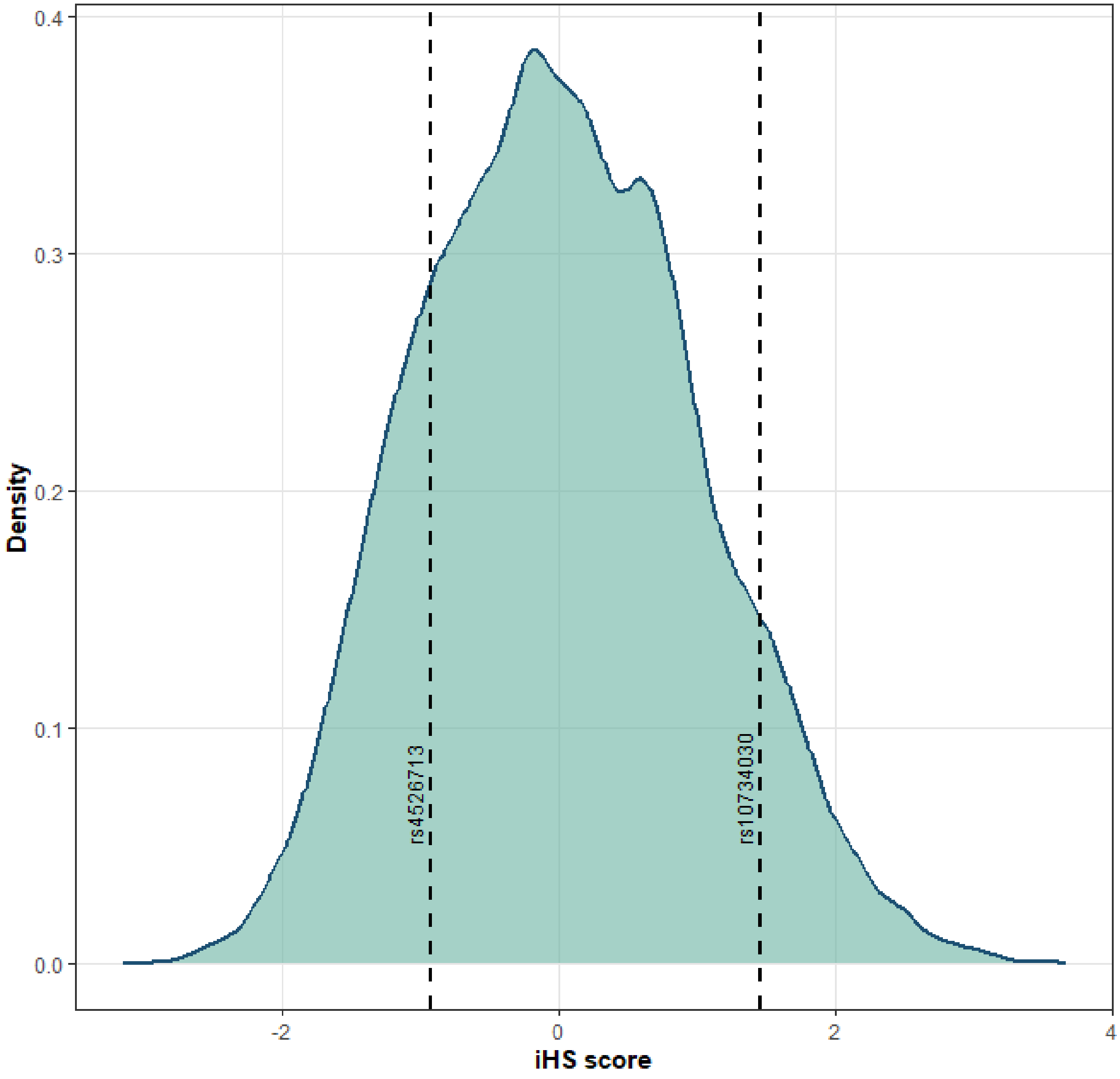


*Figure S6: iHS score distribution for SNPs within chromosome 10*

Distribution of the integrated haplotype homozygosity scores (iHS) computed using the *rehh* R package (v.3.2.3) for all chromosome 10 SNPs (n=36,182). Density was estimated using kernel smoothing and visualized with ggplot2 (v. 3.5.1). The distribution is centered around zero under the neutrality hypothesis and is expected to deviate under selection. The dashed lines indicate the iHS scores for the *PLXDC2* rs10734030 SNP (iHS=1.45) and the *ZNF518A/BLNK* rs4526713 SNP (iHS=-0.93). Both values suggest a modest excess of haplotype homozygosity around our SNPs of interest, but did not reach the significance threshold (|iHS|>2) for identifying recent selective pressure.

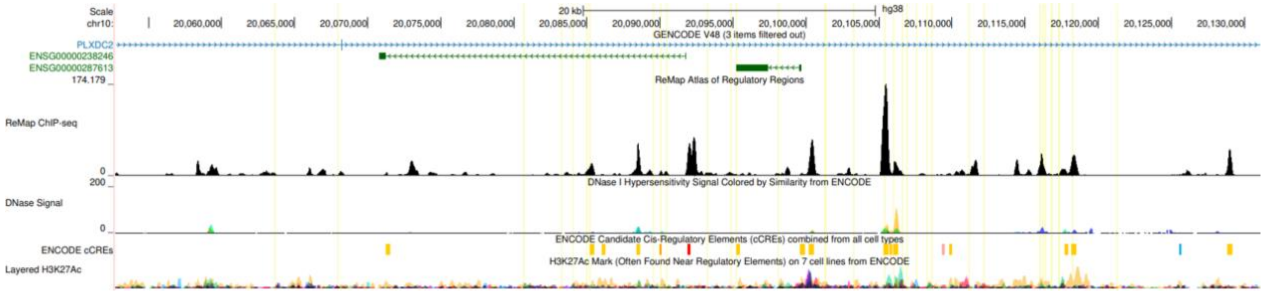


*Figure S7: UCSC Genome Browser zoomed-in view of the PLXDC2 locus (chr10:20,060,000–20,130,000, hg38).*

Ensembl gene annotations are shown at the top, with *PLXDC2* indicated in blue. The ReMap ChIP-seq peaks (black) mark transcription factor binding sites. DNase I hypersensitivity signals map regulatory elements by identifying open chromatin regions. ENCODE candidate cis-regulatory elements (cCREs) are color-coded: promoter-like (red) and enhancer-like (orange). H3K27ac ChIP-seq data (multi-colored peaks) indicate active regulatory regions across multiple ENCODE cell lines. Interestingly, several SNPs in linkage disequilibrium with the rs10734030 top SNP (vertical yellow lines) lie within open and active chromatin sites with reported enhancers, confirming the likely regulatory role of this entire region associated with CLAD.

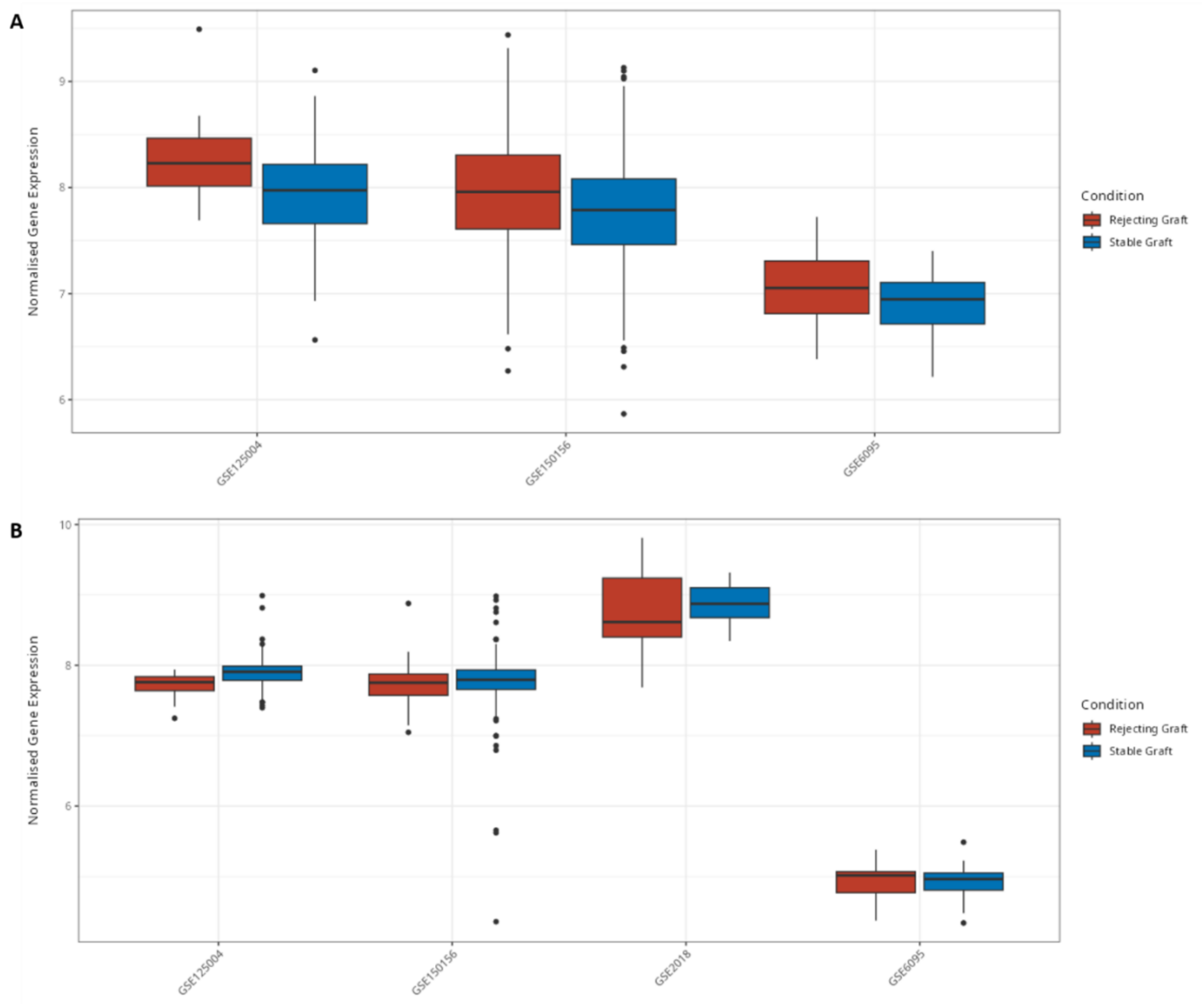


*Figure S8: Transcriptomic results for BLNK (A) and ZNF518A (B) normalized gene expressions in lung allograft rejection*

Median values, interquartile ranges, and overall variability are represented in these boxplots extracted from the PROMAD resource(3).

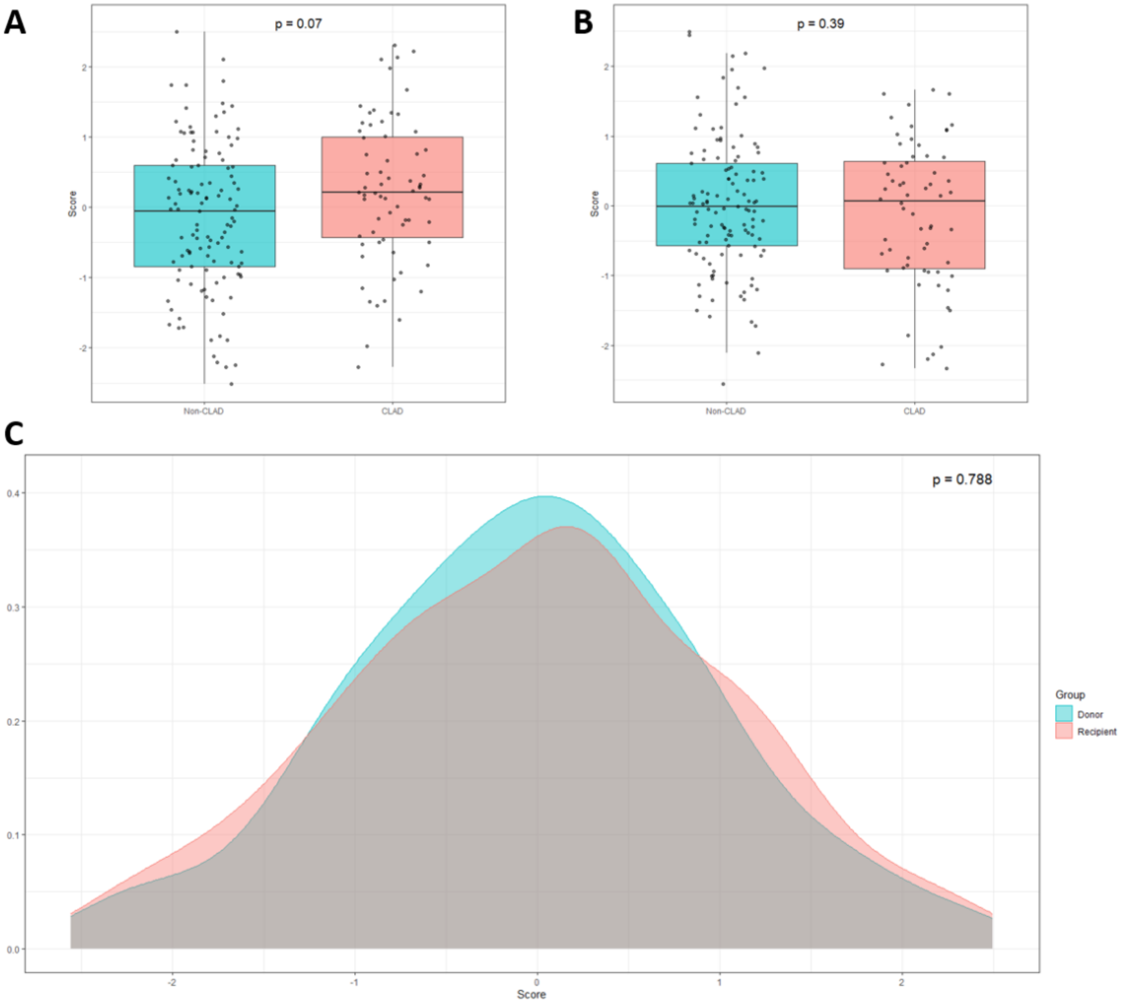


*Figure S9: Distribution of the lung function genetic risk score in the GenCOLT cohort.*

We computed the recently published multi-ancestry pulmonary function GRS(4) in the donors and recipients from the GenCOLT cohort. We compared the recipients (**A**) and donors (**B**) GRS distribution in CLAD cases vs. non-CLAD controls. We also assessed the distribution of the lung function GRS between GenCOLT recipients and donors (**C**). The lung function GRS was not associated with CLAD risk and did not discriminate end-stage lung disease cases (recipients) from controls (donors) in our cohort.

### Tables

*Table S1: Association analysis with CLAD for allelic and epitopic HLA mismatches across all class I (HLA-A, HLA-B and HLA-C), class II (HLA-DRB1 and HLA-DQB1), or overall class I+II genes (total).*

Logistic regression models assessing association with CLAD were corrected for age, sex, genetic ancestry (five first principal components), and underlying initial disease.

| | Allelic | | | Epitopic | | |
|---|---|---|---|---|---|---|
| | β | IC 95% | p-value | β | IC 95% | p-value |
| **Mismatch class I** | 0.21 | -0.13 - 0.58 | 0.24 | 0.06 | 0.02 - 0.1 | **0.004** |
| **Mismatch class II** | 0.01 | -0.4 - 0.4 | 0.97 | 0.01 | -0.03 - 0.05 | 0.67 |
| **Total mismatch** | 0.11 | -0.12 - 0.34 | 0.36 | 0.03 | 0.0 - 0.06 | **0.026** |

*Table S2: Description of the recipient's SNPs significantly associated with CLAD (FDR<5%)*

The frequency in the CLAD and non-CLAD groups is shown for the A2 allele.

| Chr | SNP | Gene | A1 | A2 | CLAD frequency | Non-CLAD frequency | Odds Ratio | Lower confidence interval | Upper confidence interval | P-value | FDR q-value |
|---|---|---|---|---|---|---|---|---|---|---|---|
| 10 | rs10734030 | *PLXDC2* | C | T | 0.385 | 0.663 | 0.346 | 0.229 | 0.523 | 5.053e-07 | 0.017 |
| 10 | rs4748637 | *PLXDC2* | T | C | 0.386 | 0.663 | 0.346 | 0.229 | 0.524 | 5.227e-07 | 0.017 |
| 10 | rs7098330 | *PLXDC2* | G | T | 0.386 | 0.662 | 0.347 | 0.229 | 0.525 | 5.443e-07 | 0.017 |
| 10 | rs7918203 | *PLXDC2* | A | G | 0.393 | 0.663 | 0.352 | 0.232 | 0.534 | 9.221e-07 | 0.017 |
| 10 | rs7917917 | *PLXDC2* | A | G | 0.393 | 0.663 | 0.352 | 0.232 | 0.535 | 9.338e-07 | 0.017 |
| 10 | rs10764200 | *PLXDC2* | A | G | 0.350 | 0.624 | 0.351 | 0.231 | 0.533 | 9.339e-07 | 0.017 |
| 10 | rs7085207 | *PLXDC2* | A | G | 0.393 | 0.663 | 0.353 | 0.232 | 0.535 | 9.526e-07 | 0.017 |
| 10 | rs4526713 | *ZNF518A* | C | T | 0.659 | 0.382 | 2.834 | 1.867 | 4.301 | 9.864e-07 | 0.017 |
| 10 | rs7071511 | *PLXDC2* | A | C | 0.394 | 0.663 | 0.352 | 0.232 | 0.535 | 9.923e-07 | 0.017 |
| 10 | rs11599170 | *ENTPD1-AS1* | A | G | 0.664 | 0.387 | 2.813 | 1.856 | 4.264 | 1.093e-06 | 0.017 |
| 10 | rs11188622 | *ENTPD1-AS1* | G | A | 0.660 | 0.385 | 2.825 | 1.859 | 4.292 | 1.144e-06 | 0.017 |
| 10 | rs11595809 | *ENTPD1-AS1* | C | T | 0.660 | 0.385 | 2.824 | 1.858 | 4.290 | 1.151e-06 | 0.017 |
| 10 | rs6482085 | *PLXDC2* | A | C | 0.358 | 0.632 | 0.356 | 0.234 | 0.539 | 1.162e-06 | 0.017 |
| 10 | rs138218133 | *ZNF518A* | AT | A | 0.655 | 0.382 | 2.767 | 1.827 | 4.193 | 1.567e-06 | 0.017 |
| 10 | rs34999669 | *ZNF518A* | G | T | 0.655 | 0.382 | 2.767 | 1.827 | 4.193 | 1.569e-06 | 0.017 |
| 10 | rs11598837 | *ZNF518A* | G | A | 0.655 | 0.382 | 2.767 | 1.827 | 4.192 | 1.569e-06 | 0.017 |
| 10 | rs10491070 | *BLNK* | T | C | 0.651 | 0.378 | 2.782 | 1.832 | 4.223 | 1.574e-06 | 0.017 |
| 10 | rs10509701 | *ZNF518A* | G | A | 0.654 | 0.381 | 2.759 | 1.822 | 4.179 | 1.647e-06 | 0.017 |
| 10 | rs34302998 | *ZNF518A* | CT | C | 0.654 | 0.381 | 2.758 | 1.821 | 4.177 | 1.665e-06 | 0.017 |
| 10 | rs12761705 | *ZNF518A* | C | T | 0.654 | 0.381 | 2.757 | 1.821 | 4.176 | 1.672e-06 | 0.017 |
| 10 | rs12354644 | *ZNF518A* | A | C | 0.654 | 0.381 | 2.755 | 1.819 | 4.172 | 1.699e-06 | 0.017 |
| 10 | rs11188632 | *ZNF518A* | C | T | 0.653 | 0.381 | 2.742 | 1.812 | 4.149 | 1.839e-06 | 0.017 |
| 10 | rs11188631 | *ZNF518A* | G | A | 0.653 | 0.381 | 2.740 | 1.811 | 4.147 | 1.854e-06 | 0.017 |
| 10 | rs7896077 | *ZNF518A* | T | C | 0.653 | 0.381 | 2.740 | 1.811 | 4.147 | 1.859e-06 | 0.017 |
| 10 | rs11598546 | *ZNF518A* | A | C | 0.653 | 0.381 | 2.740 | 1.810 | 4.146 | 1.866e-06 | 0.017 |

| | | | | | | | | | | | |
|---|---|---|---|---|---|---|---|---|---|---|---|
| 10 | rs11593049 | *ZNF518A* | G | A | 0.653 | 0.381 | 2.738 | 1.809 | 4.143 | 1.891e-06 | 0.017 |
| 10 | rs60450792 | *ZNF518A* | T | C | 0.653 | 0.381 | 2.736 | 1.808 | 4.141 | 1.916e-06 | 0.017 |
| 10 | rs10509700 | *ZNF518A* | G | C | 0.653 | 0.382 | 2.728 | 1.803 | 4.127 | 2.034e-06 | 0.017 |
| 10 | rs34673323 | *ZNF518A* | TC | T | 0.653 | 0.382 | 2.728 | 1.803 | 4.127 | 2.034e-06 | 0.017 |
| 10 | rs10882723 | *ZNF518A* | A | G | 0.653 | 0.382 | 2.727 | 1.803 | 4.126 | 2.046e-06 | 0.017 |
| 10 | rs7902212 | *PLXDC2* | T | C | 0.378 | 0.639 | 0.371 | 0.246 | 0.560 | 2.377e-06 | 0.019 |
| 10 | rs7089618 | *PLXDC2* | C | T | 0.343 | 0.610 | 0.372 | 0.246 | 0.562 | 2.636e-06 | 0.020 |
| 10 | rs7916674 | *PLXDC2* | A | T | 0.344 | 0.608 | 0.369 | 0.243 | 0.559 | 2.673e-06 | 0.020 |
| 10 | rs10764201 | *PLXDC2* | C | T | 0.344 | 0.608 | 0.369 | 0.243 | 0.560 | 2.766e-06 | 0.020 |
| 10 | rs2125060 | *ENTPD1-AS1* | A | C | 0.682 | 0.417 | 2.700 | 1.781 | 4.095 | 2.927e-06 | 0.021 |
| 10 | rs34211982 | *PLXDC2* | T | TG | 0.383 | 0.642 | 0.377 | 0.249 | 0.571 | 4.014e-06 | 0.028 |
| 10 | rs2255452 | *PLXDC2* | C | T | 0.636 | 0.384 | 2.612 | 1.732 | 3.937 | 4.569e-06 | 0.030 |
| 10 | rs2461945 | *PLXDC2* | G | A | 0.636 | 0.384 | 2.611 | 1.732 | 3.938 | 4.638e-06 | 0.030 |
| 10 | rs2460578 | *PLXDC2* | G | T | 0.635 | 0.384 | 2.608 | 1.729 | 3.934 | 4.899e-06 | 0.031 |
| 10 | rs7096382 | *PLXDC2* | A | G | 0.356 | 0.608 | 0.385 | 0.255 | 0.580 | 5.114e-06 | 0.031 |
| 10 | rs7090580 | *PLXDC2* | T | G | 0.357 | 0.609 | 0.385 | 0.255 | 0.580 | 5.141e-06 | 0.031 |
| 10 | rs2461944 | *PLXDC2* | T | C | 0.636 | 0.388 | 2.581 | 1.708 | 3.901 | 6.756e-06 | 0.040 |
| 10 | rs2252516 | *PLXDC2* | C | A | 0.635 | 0.388 | 2.579 | 1.706 | 3.898 | 7.064e-06 | 0.040 |
| 10 | rs2460592 | *PLXDC2* | G | A | 0.634 | 0.388 | 2.576 | 1.703 | 3.896 | 7.374e-06 | 0.040 |
| 10 | rs7090610 | *PLXDC2* | T | C | 0.357 | 0.604 | 0.389 | 0.258 | 0.588 | 7.381e-06 | 0.040 |
| 10 | rs2262890 | *PLXDC2* | G | A | 0.629 | 0.384 | 2.566 | 1.699 | 3.877 | 7.584e-06 | 0.040 |
| 10 | rs2461938 | *PLXDC2* | A | C | 0.629 | 0.384 | 2.565 | 1.698 | 3.875 | 7.625e-06 | 0.040 |
| 10 | rs2460599 | *PLXDC2* | A | T | 0.608 | 0.365 | 2.552 | 1.688 | 3.857 | 8.843e-06 | 0.046 |
| 10 | rs12265061 | *LINC02624* | A | T | 0.216 | 0.067 | 4.555 | 2.327 | 8.915 | 9.621e-06 | 0.046 |
| 10 | rs10491069 | *BLNK* | T | G | 0.646 | 0.396 | 2.563 | 1.689 | 3.889 | 9.728e-06 | 0.046 |
| 10 | rs2461946 | *PLXDC2* | T | C | 0.644 | 0.400 | 2.530 | 1.676 | 3.819 | 9.878e-06 | 0.046 |
| 10 | rs2250374 | *PLXDC2* | T | C | 0.644 | 0.400 | 2.530 | 1.676 | 3.819 | 9.888e-06 | 0.046 |
| 10 | rs2460590 | *PLXDC2* | G | A | 0.629 | 0.388 | 2.537 | 1.676 | 3.841 | 1.078e-05 | 0.046 |
| 10 | rs2460585 | *PLXDC2* | C | G | 0.629 | 0.388 | 2.536 | 1.675 | 3.839 | 1.091e-05 | 0.046 |
| 10 | rs2461937 | *PLXDC2* | T | C | 0.629 | 0.388 | 2.536 | 1.675 | 3.839 | 1.092e-05 | 0.046 |
| 10 | rs2461939 | *PLXDC2* | C | T | 0.629 | 0.388 | 2.535 | 1.675 | 3.838 | 1.094e-05 | 0.046 |
| 10 | rs2461940 | *PLXDC2* | T | C | 0.629 | 0.388 | 2.535 | 1.675 | 3.838 | 1.095e-05 | 0.046 |
| 10 | rs2461942 | *PLXDC2* | C | G | 0.629 | 0.388 | 2.535 | 1.675 | 3.838 | 1.098e-05 | 0.046 |

| | | | | | | | | | | | |
|---|---|---|---|---|---|---|---|---|---|---|---|
| 10 | rs2460583 | *PLXDC2* | G | A | 0.629 | 0.388 | 2.536 | 1.675 | 3.839 | 1.100e-05 | 0.046 |
| 10 | rs2461936 | *PLXDC2* | A | C | 0.629 | 0.388 | 2.535 | 1.675 | 3.839 | 1.102e-05 | 0.046 |
| 10 | rs2461941 | *PLXDC2* | C | T | 0.634 | 0.391 | 2.513 | 1.666 | 3.790 | 1.118e-05 | 0.046 |
| 10 | rs7912564 | *PLXDC2* | C | G | 0.274 | 0.508 | 0.381 | 0.247 | 0.586 | 1.172e-05 | 0.047 |
| 10 | rs12777653 | *PLXDC2* | C | G | 0.560 | 0.325 | 2.578 | 1.687 | 3.940 | 1.205e-05 | 0.048 |
| 10 | rs147898547 | *PLXDC2* | TA | T | 0.559 | 0.325 | 2.574 | 1.683 | 3.936 | 1.280e-05 | 0.049 |
| 10 | rs79952085 | *PLXDC2* | A | T | 0.559 | 0.325 | 2.574 | 1.683 | 3.936 | 1.280e-05 | 0.049 |

*Table S3: Logistic regression models assessing the association between SNP mismatch scores and CLAD risk*

Akin to what was proposed in kidney transplantation(5,6), the strategy was to summarize the overall non-HLA SNP mismatch burden within a donor-recipient pair by focusing on alleles coding for protein epitopes expressed by the donor organ that the recipient's immune system could recognize as non-self. We investigated 4 different SNP subsets to compute the sum of mismatches: (1) all non-synonymous SNPs, (2) all non-synonymous SNPs from genes previously associated with lung function in a recent GWAS(4), (3) all non-synonymous SNPs from genes previously associated with a lung disease in the GWAS Catalog(7), and (4) all exonic SNPs. Logistic regression models were used to evaluate the association between these different SNP mismatch scores and the CLAD risk while adjusting for age, sex, genetic ancestry (first five principal components), and the underlying initial disease. Negative β values indicate a trend towards lower CLAD risk with higher mismatch scores, although none of the associations were statistically significant.

| Donor-recipient SNP mismatch burden scores | Number of SNPs | β | p-value |
|---|---|---|---|
| **Non-synonymous SNP mismatch score** | 15,192 | -8.4e-6 | 0.14 |
| **Lung-function specific non-synonymous SNP mismatch score(4)** | 799 | -4.5e-3 | 0.24 |
| **Lung-diseases specific non-synonymous SNP mismatch score(7)** | 1,475 | -4.4e-5 | 0.29 |
| **Exonic SNP mismatch score** | 1,031,760 | -5.1e-5 | 0.28 |

### Supplementary Methods

#### *GenCOLT biobank*

The study includes all adult patients (aged 18 and older) who survived at least three months after LT, given that the recipient provided consent for genetic analyses and that DNA samples were available for both the donor and the recipient. The collection of samples from deceased donors for scientific research has been authorized by the French biomedical agency (*Agence de Biomédecine*, PFS09-003), and a protocol has been established to verify whether the donor objected to the use of their organs or tissues for research and to provide information to their relatives. The GenCOLT cohort is located and managed by the biological resource center (*Centre des Ressources Biologiques)* at the Nantes University Hospital(8).

#### *Genotyping*

For each individual, genomic DNA was isolated from whole blood. SNP genotyping was performed following the manufacturer recommendations using the Axiom PMRA (Precision Medicine Research Array) chips (ThermoFisher, Waltham MA, USA), which cover 902,560 genetic variants including those found in the *HLA* and *KIR* polymorphic genomic regions, as well as other relevant genes for research in cancer and immunology. We followed the Axiom 2.0 ThermoFisher standardized protocols and guidelines during the genotyping process.

#### *Quality control steps*

To ensure the reliability and integrity of the genomic dataset, we applied a comprehensive multi-step quality control (QC) procedure. Initially, a technological QC step was conducted using the Axiom Analysis Suite (AxAS) to retain only high-quality individuals, plates, and variants. For individual-level filtering, we required a DishQC score >0.82 to confirm the clear separation of AT and GC fluorescence signals from background noise on nonpolymorphic probes, and a genotype call rate >97%. Plates were retained if their call rate exceeded 98.5%, and genetic variants with a call rate above 95% were included. In addition, SNPs were assessed for clustering quality using Fisher's linear discrimination (>3.6) and clear differentiation between homozygous and heterozygous genotypes (heterozygosity >95%). Following this technological QC, 852,344 SNPs and 387 sample pairs (n = 775 individuals) were retained.

Further QC steps were performed using PLINK(9) to reduce the risk of genotyping errors. Individuals with >2% missing data were flagged, and relatedness among samples was examined; no exclusions were made at this stage. SNPs with >2% missingness or deviating significantly from the Hardy-Weinberg equilibrium ($p < 10^{-6}$) were removed.

To address missing genotypes, we performed SNP imputation using the TOPMed imputation server(10) and its corresponding reference panel. For *HLA* allele imputation, the HIBAG algorithm(11) was used with a global reference panel comprising multiethnic samples from the 1000 Genomes Project(12). In both cases, only SNPs and *HLA* alleles with high imputation accuracy ($r^2 > 0.8$) were retained for downstream analyses.

#### *SNP and HLA association testing*

Each genotyped and imputed SNP from the donors or from the recipients were tested for association with CLAD using SAIGE (v.0.45.)(13). Multivariable logistic regression models were adjusted with donors' (resp. recipients') age and sex, the underlying initial pulmonary disease and population structure and relatedness (genetic ancestry) calculated by the genomic relationship matrix (GRM). Donor-recipient mismatches were tested for association with CLAD using generalized linear models (GLMs) implemented in R and the following covariates: donor and recipient age and sex, the first five principal components (PCs) for genetic ancestry, and the underlying initial disease. We accounted for multiple testing by computing the Benjamin-Hochberg false discovery rate (FDR, q-value), which is a relevant and robust multiple testing correction method increasing true positive discovery(14). SNPs with an FDR q-value <5% were considered significant.

Similarly, *HLA* alleles were tested for association with CLAD using multivariable logistic regression models corrected for age, sex, genetic ancestry (5 first PCs) and the initial disease. The dominant genetic model of inheritance was tested for *HLA* immunogenetic parameters. Allelic and epitopic HLA mismatch scores were also evaluated using multivariable logistic regression models adjusted for the same covariates as above.

Manhattan and Quantile-Quantile (QQ) plots were generated with R (v.4.4.2) to present the association results with CMplot (v. 4.5.1)(15). The zoomed-in regional association plots were generated with locuszoom (v. 0.3.8)(16). The statistical power was evaluated according to several ranges of odds-ratio and minor allele frequency (MAF) by genpwr (v1.0.4)(1).

#### *Polygenic risk scores computation*

We first computed the recently published multi-ancestry genetic risk score (GRS) constructed from lung function (FEV1/FVC ratio) GWAS and associated with chronic obstructive pulmonary disease (COPD) risk, by using their published list of SNPs and weights(4). We then fitted a logistic regression model to test the association between the lung function GRS in donors or recipients and the CLAD/non-CLAD status while adjusting for age, age squared, sex, height, underlying disease, and the first five principal components.

Akin to what was proposed in kidney transplantation(5,6), we also computed several non-HLA mismatch scores by focusing on non-synonymous mismatches that could represent potential immunogenic antigens expressed by the donor organ and recognize as non-self by the recipient. The underlying hypothesis is that each donor-recipient pair might carry different mismatch combinations and that the CLAD risk would increase with a higher mismatch load. In addition, as non-HLA mismatch scores sum the contribution of thousands of mismatches across the genome into one burden score, this strategy also decreases the need for high sample sizes for statistical power discovery. We investigated 4 different SNP subsets (MAF≥1%) to compute the mismatch scores: (1) all non-synonymous SNPs, (2) all non-synonymous SNPs from genes previously associated with lung function in a recent GWAS(4), (3) all non-synonymous SNPs from genes previously associated with a lung disease in the GWAS Catalog(7), and (4) all exonic SNPs. Logistic regression models were used to evaluate the association between each of these scores and the CLAD risk while adjusting for age, sex, genetic ancestry (first five principal components), and the underlying initial disease.

#### *Lung sample preparation for scRNAseq*

Lungs were cut in 1x1cm pieces and put in digestion media DMEM, collagenase 1mg/mL, DNase Deoxyribonuclease I from bovine pancreas 0.1mg/mL, penicillin (100 UI/mL), streptomycin (100 µg/mL) (P/S) and amphotericin B 0,25µg/mL) for 45 min at 37°C. Cells were filtered through a 70-µm strainer then pelleted (360g, 10 min, 4°C) and resuspended in 10% fetal calf serum DMEM three times. Cells were then resuspended and incubated in red blood cell lysis buffer for 5 min at room temperature after which the buffer was inactivated by adding 50 mL of cold phosphate buffer saline (PBS) and pelleted again. Cells from freshly dissociated lung explants were then stored at -80°C in fetal bovine serum with 10% DMSO until use. Cells were thawed and sorting for living cells was performed by flow cytometry (ARIA III, BD Biosciences) using DAPI viability marker. The cells were suspended using Chromium 10X chip (10X Genomics Technology) and given a unique identifier. RT-PCR was performed using a GEM-X Single cell 3' v4 library according to the supplier's instructions. Sequencing of the resulting cDNAs was performed on a flow cell S2 of a Nova-Seq 6000 instrument (Illumina) on the GenoBiRD platform (IRS-UN, CHU Nantes). Doublets and cells expressing over 15% of mitochondrial RNA were excluded.

#### *scRNAseq data analysis*

We obtained the gene count matrix at the individual cell level using Cellranger (v7.0.1) against the hg38 human reference genome. Downstream analyses were performed within the Seurat (v5.2.1) framework on R (v4.4.2). Senescent, doublet, or cells with low sequencing depth were discarded. We used Harmony (v1.2.3) to integrate data generated from different batches. We applied UMAP dimensional reduction and clustering steps to the integrated dataset, and cell cluster identification was carried out using the Azimuth (v0.5.0) package. Finally, we analyzed differential gene expression between groups of interest (cell cluster or phenotype) on unaggregated expression profiles using a Wilcoxon test.

### Supplementary Results

#### *Demographics and clinical characteristics*

Overall, the CLAD and non-CLAD subgroups were not different ($p \geq 0.05$) regarding the recipient's and donor's demographic characteristics (including sex, type of procedure, serology, treatment, see Tables 1 and 2), except for age and follow-up time. Indeed, the recipients and donors from the CLAD group were on average older than in the non-CLAD group: 51 vs. 45 years-old for the recipients (p=0.007) and 50 vs. 42 years-old for the donors (p<0.001). As expected, the follow-up time was shorter for individuals experiencing CLAD (5.14 years vs. 6.60 years, p<0.001). Although not significant, the CLAD group was more frequently transplanted for chronic obstructive pulmonary disease (COPD) than the non-CLAD group (60% vs. 40%). Finally, there was no significant difference for donor's causes of death (p=0.12), smoking habit (p=0.3) and $PaO_2/FiO_2$ ratio (p=0.2).

#### *Selection signature*

Interrogating the 1000 Genomes public database, we noticed an interesting pattern of global frequency distribution for the *PLXDC2* rs10734030-C allele from barely 3% in Sub-Saharan Africa to 0% in East-Asia and up to 35-45% in Europe, which could reflect a genetic drift or an evolutionary selective pressure after the out-of-Africa event. The frequency distribution of the rs4526713-T allele was more homogeneous across continents, ranging from 18% in Sub-Saharan Africa, to 27% in South-Asia and 38-48% in Europe, even if the frequency was only 3% in East-Asia. We therefore set out to explore the recent selection signature around our two top SNPs using the population differentiation $F_{ST}$ index and the integrated haplotype score (iHS) (for detecting ongoing or recent selective sweeps).

The $F_{ST}$ index distribution across the human genome was centered on $F_{ST}=0$ indicating that most SNPs cannot differentiate between two continental populations (Figure S5). However, the $F_{ST}$ index for the *PLXDC2* rs10734030 SNP was 0.37 in Europeans vs. the other populations and in the extreme top 1$^{st}$

percentile of the distribution, which indicates potential local adaptation(17),(18). The $F_{ST}$ index value for the *ZNF518A/BLNK* rs4526713 SNP was 0.16 in Europeans (<5% of the distribution), hence not emphasizing any selection event.

The iHS score distribution was centered around 0, indicating no widespread signal of recent positive selection (Figure S6). However, the iHS values for the *PLXDC2* rs10734030 and *ZNF518A/BLNK* rs4526713 SNPs were 1.45 and -0.93, respectively. These values suggest a modest excess of haplotype homozygosity around one allele at each locus, consistent with a potential ongoing or recent selective sweep, although they fall below the commonly used threshold (|iHS|>2) for identifying strong candidates of recent selection.

#### *Polygenic risk scores*

We first investigated if the recent multi-ancestry lung function GRS was associated with CLAD risk in our cohort. Overall, 89% (910/1,020) SNPs included in the original GRS were present in our dataset. Neither the recipient nor the donor GRS were associated with CLAD (p=0.07 and p=0.39, respectively, Figures S8A-B). In addition, the GRS distribution was similar in recipients, who all experienced lung failure before transplantation, and in donors who did not (p=0.79, Figure S8C).

Second, we investigated if an overall non-HLA mismatch score could capture the overall burden of potential immunogenic antigens presented by the donor to the recipient as previously proposed in kidney transplantation. None of the score configurations we tested (all non-synonymous mismatches, restricting to lung function genes, or to lung disease genes) were associated with CLAD risk in our cohort (p>0.1, Table S3).
